# Gut Microbial Dysbiosis and Elevated Mucosal Inflammation in Severe Acute Malnutrition: A Case-Control Study from India

**DOI:** 10.1101/2025.11.04.25339370

**Authors:** Parth Sarin, Harshitha Yadav, Raj Ojha, Santosh Kumar Banjara, Sakshi Rai, Manoj Kumar, Hemant Mahajan, Samradhi Singh, JJ Babu Geddam, Aruna Reddy V, G Vijayalaxmi, Jhansi Reddy N, Nikhita Boda, Sourav Sen Gupta, Devraj J Parsannanavar

**Affiliations:** Department of Clinical Epidemiology, ICMR-National Institute of Nutrition, India; Anne Spencer Daves College of Education, Health, and Human Sciences, Florida State University, USA; Department of Maternal and Child Health, ICMR-National Institute of Nutrition, India; Department of Microbiology, ICMR- National Institute of Research in Environmental Health, Bhopal, India; Department of Pathology and Laboratory Medicine, Emory University School, United States of America

**Keywords:** Gut microbiota, Nutrition, Paediatric health, Inflammation, Socioeconomic status

## Abstract

Severe Acute Malnutrition (SAM) in early days of life has been a concerning global health challenge, contributing to high child mortality and morbidity. The mechanistic role of the gut microbiota in its pathophysiology remains incompletely understood. In this case-control study we profiled gut-microbiota using 16S-rRNA sequencing for V3-V4 region for 115 SAM children (52 SAM and 63 Healthy) aged 6-59 months. Inflammatory markers along with fecal sIgA levels were analysed and correlated with clinical parameters, and dietary intake of these children. Alpha diversity has revealed a significant difference in ChaoI index indicating reduced richness in SAM group (p<0.001). Children with SAM have also shown reduced abundance of beneficial groups such as Blautia, Akkermansia, and Ruminococcus gnavus, with an expansion of opportunistic groups like Enterococcus, Staphylococcus, and Turicibacter. Inflammatory markers show elevated levels of inflammation in SAM children when compared to healthy controls. Reduced fecal sIgA levels in SAM group have been noted, suggesting an impaired immune response. Male children were at higher risk of inflammation and immune dysfunction. Our findings delineate gut-microbial disruption as a central feature of childhood SAM. Targeting these specific taxa and their associated metabolic pathways can help in tailoring future interventions.

## 1. INTRODUCTION

Nutrition is fundamental to child growth, immune development, and long-term health outcomes. Undernutrition, resulting from insufficient intake of balanced food groups, during critical developmental periods, including fetal life, infancy, and early childhood can severely impair child’s growth, development, and overall quality of life (1,2). The most extreme and visible manifestation of undernutrition is **Severe Acute Malnutrition (SAM)**, which is characterized by rapid weight loss, muscle wasting, poor nutrient absorption, and physiological alterations such as gut inflammation, environmental enteric dysfunction (EED), and compromised intestinal barrier function (3).

SAM in children who are 6–59 months of age, is defined as the presence of oedema of both feet or severe wasting (weight-for-height/length <-3SD or mid-upper arm circumference < 115 mm). Globally, SAM affects approximately 19 million children under five and contributes to an estimated 400,000 child deaths annually(4,5).

The prevalence of SAM among Indian preschool children has shown a marginal increase over the past two decades, rising from 6.6% in the National Family Health Survey-3 (2005–06) to 7.7% in NFHS-5 (2019–21). SAM poses a **9–11× higher risk of mortality** compared to healthy children. India contributes nearly one-third of the global burden of undernutrition and has the highest number of children with severe wasting worldwide. Children affected by SAM are at elevated risk of recurrent infections, delayed development, and premature death, making it a critical public health concern requiring urgent intervention(6).

While, inadequate diet and socioeconomic disparities are primary contributors to SAM(7,8), emerging evidence suggest complex interaction of gut microbiota, inflammation and nutrient absorption. SAM is often accompanied with persistent diarrhoea(9,10), altered morphology and function of the gut (villus blunting), and a dysregulated immune response. Combined effect of these factors lead to poor nutrient absorption and sustained malnutrition(11).

A key player in this cycle is gut microbiota dysbiosis, characterized by reduced microbial diversity, loss of beneficial commensals, and an overrepresentation of pathogenic and pro-inflammatory taxa. Various studies also highlight decreased gut microbial diversity in SAM children when compared to their healthy counterparts, highlighting its role in disease pathophysiology(12,13).

Recent study found that Children with SAM were identified to have an immature gut microbiota when compared to healthy children of similar age (14). A study conducted in Bangladesh also revealed a substantial increase in the abundance of the phylum Proteobacteria, including pathogenic genera such as Klebsiella and Escherichia, and a decrease in the abundance of the phylum Bacteroidetes in malnourished children compared to healthy children (15). Another study in Indonesia revealed a significant difference in the abundance of *Firmicutes*, *Bacteroidetes*, *Actinobacteria*, *Proteobacteria* and *Verrucomicrobia* between undernourished and healthy children (12). These studies highlight the critical aspect of gut microbiota in undernourished children.

Despite advancements in therapeutic approaches such as ready-to-use therapeutic foods (RUTF) and fortified supplements, current treatment strategies largely overlook gut microbiota dimension of SAM(16). Addressing gut dysbiosis may represent a promising adjunct to nutritional rehabilitation, yet microbial targets remain poorly characterized in the Indian context.

This study aims to fill that gap by examining the **gut microbial profiles of Indian preschool children with SAM** compared to healthy counterparts. By exploring the complex interactions between gut microbial communities, inflammation, and the nutritional status of children, the study seeks to identify specific microbial patterns associated with SAM. This work will enhance our understanding of the microbiota-host-nutrition axis and inform microbiota-based or integrative therapeutic strategies for childhood malnutrition.

## 2. MATERIALS AND METHODS

### 2.1 Ethical Statement

The ethical approval for the project was obtained from the Institutional Ethics Committee (IEC approval No. 05/II/2018, dated 01/11/2018) and the Integrated Child Development Service (ICDS). Written informed consent was obtained from parents/guardians of the subjects recruited for the study.

### 2.2 Study design and selection of subjects

In this case-control study, children aged 6-59 months were selected based on specific inclusion and exclusion criteria as mentioned in the CONSORT diagram. The CONSORT diagram underlines the number of Anganwadi and children screened before recruitment and final participants taken for analysis (Figure S4). A total of 129 children from all ethnic and social groups were recruited for the study from Anganwadi centres in undernutrition-prone divisions, as reported by the Integrated Child Development Services (ICDS) in Hyderabad, Telangana. Detailed sociodemographic, clinical, and dietary information for each subject was recorded according to a structured proforma and questionnaires(Supplementary S2). Children with any chromosomal anomalies, history of diarrhoea, upper and lower respiratory illness or any other serious ailment requiring hospital admission within at least four weeks before faecal sample collection were excluded. Additionally, children who were taking any antibiotics, antacids, probiotics, prebiotics, or medications containing alcohol within at least four weeks before faecal sample collection were excluded.

### 2.3 Anthropometric measurement

The children’s height, weight and mid-upper arm circumference (MUAC) were measured at Anganwadi centres with the help of Anganwadi teachers and staff. Weight, precision to the nearest 0.1 kg, was measured using a SECA digital weighing scale (Seca 813) while the participant stood barefoot and wore minimal clothing in the centre of the scale. Height, precision to the nearest 0.1 cm, was measured using a SECA height rod (Seca 213 stadiometer) while the participant stood straight with head, shoulders, buttocks and heels touching the flat surface, looking straight ahead and arms at the sides. Recumbent length (Baby length) was measured for the children under 24 months using an infantometer (Seca 416). The MUAC of the non-dominant arm(left arm) was measured using a measuring tape at the midpoint between the tip of the shoulder and the tip of the elbow.

Based on anthropometric measurements, children whose weight-for-height z-score (WHZ) was less than −3, the median of the WHO child growth standards, were grouped into the SAM category and those between −1 and +1 of the medians of the WHO child growth standards were grouped into the healthy category.

### 2.4 Dietary and sociodemographic data collection

Data of the recruited subjects(n=117) was collected using three different questionnaires: general sociodemographic information, 24-hour dietary recall, and Food Frequency questionnaire(FFQ) from the mothers of recruited children and teachers at the Anganwadi centres. Detailed demographic data, birth history, sanitation and hygiene practices, feeding and dietary practices, and the subject’s illness status were collected with a pre-designed questionnaire using the KoBocollect application.

#### Diet data collection

A single 24-Hour Dietary Recall (24-HDR) and FFQ survey was conducted among children’s caretakers or mothers by a field investigator. Detailed dietary intake data of the previous day were collected using a standardised methodology, i.e., detailed information on foods that were cooked and served at home, along with consumption of outside foods. Their mealtime, list of recipes, and ingredients were recorded. After listing the ingredients against each recipe, amounts used in the recipes were weighed using a calibrated weighing scale and noted in grams or millilitres. The equation used to calculate further raw food quantities consumed from the cooked food consumed (Supplementary S3).

The nutrient composition of the consumed food was then computed using the Indian Food Composition Tables 2017. Frequency of 131 food groups was also collected using FFQ, with consumption frequency analysed in daily, weekly, monthly and yearly.

### 2.5 Inflammatory markers measurement

Inflammatory markers were assessed in a subset of samples, including myeloperoxidase (MPO) and calprotectin (CAL), were measured in 82 samples comprising 41 age and sex-matched SAM children with 41 healthy controls, using sandwich ELISA kits (IDK; Cat. No. KR6630 and KR6927) according to the manufacturer’s protocols. Secretory IgA (sIgA) was analysed in 84 samples, including 42 SAM and 42 healthy children, following the manufacturer’s protocol (Immunochrom; Cat. No. IC6100). Neopterin (NEO) was assessed in 82 samples, using a competitive ELISA kit (IBL International; Cat. No. RE59321), following the manufacturer’s instructions, with normal saline(0.9%) used for diluting faecal samples. All assays were performed in duplicate, and mean absorbance values were used to calculate concentrations. Analyte concentrations were determined using a four-parameter logistic (4PL) standard curve.

### 2.6 Statistical analysis of diet and sociodemographic data

Statistical analyses were conducted using Stata version 15.0. Continuous variables with skewed distributions were summarised as medians with interquartile ranges and compared between groups using the Mann-Whitney U test. Categorical variables were expressed as frequencies (percentages), and differences between groups were assessed using Pearson’s Chi-Square test, with corresponding p values reported. To identify risk factors for SAM, univariable and multivariable logistic regression analyses were performed, with risks expressed as odds ratios and 95% confidence intervals. A two-tailed p value of <0.05 was considered statistically significant.

### 2.7 Faecal Sample Collection, DNA Extraction and 16S rRNA Gene Amplicon Sequencing

#### Sample Collection and DNA Extraction

We recruited 129 participants(72 healthy and 57 SAM) in the study, but could only collect samples from 115 participants (63 healthy and 52 SAM). Fresh fecal samples were collected in sterile 50 mL containers(Tarsons: Cat no.-510030), parents were instructed to not contaminate the samples with urine and store it in provided dry ice boxes.

Samples were transported in the same box maintaining the cold chain and were immediately stored at −80°C upon arrival until further processing. Sample collection date was maintained by the field staff to keep the proper record. A total of 115 samples were subjected to metagenomic analysis. DNA extractions were performed using the QIAamp PowerFecal Pro DNA Kit (Qiagen;Cat no.51804), incorporating bead-beating during lysis in the manufacturer’s recommended protocol.

The isolates were subjected to amplification of the V3–V4 hypervariable region of the 16S rRNA gene using primers 341F (CCTAYGGGRBGCASCAG) and 806R (GGACTACNNGGGTATCTAAT). The amplified products underwent library preparation using the **NEBNext® Ultra™ II DNA Library Prep Kit (New England Biolabs; Cat No. E7645)**, which included end-repair, A-tailing, and ligation with Illumina-compatible adapters. The NEBNext libraries resemble TruSeq libraries and were processed accordingly, with adapter sequences trimmed as follows: **Read 1** AGATCGGAAGAGCACACGTCTGAACTCCAGTCA; Read 2 **AGATCGGAAGAGCGTCGTGTAGGGAAAGAGTGT**. Library quality and fragment size distribution were assessed using a bioanalyzer (AATI), and concentrations were determined via quantitative PCR (qPCR). Afterwards, Equimolar amounts of purified PCR products from each sample were pooled and sequenced on the Illumina NovaSeq 6000 platform using SP flow cell with PE250 chemistry to generate ∼0.1 million non-chimeric reads.

### 2.8. Data processing and Statistical analysis

The metagenomic raw reads was demultiplexed and post quality checks using q2-demux plugin. Afterwards, denoising and chimera filtering was performed using DADA2 (17). The sequences were merged and aligned with MAFFT(18) sub-sequentially taxonomic assignment was done with sklearn naïve Bayes classifier aligning on 99% SILVA v1.3.8 database. For cluster analysis Bray-Curtis distance matrix was calculated and dimensionality reduction was performed using t-SNE. The significance in differences between the clusters was obtained by performing PERMANOVA. The differential abundance analysis was performed using the Mann-Whitney U test with Benjamini-Hochberg FDR correction, LEfSe, and MaSLin3. The correlations between the other parameters and microbial genera were performed with Spearman test and significance was marked at p-values of <0.1 and <0.05 unadjusted. Only the significantly different microbial genera in Mann-Whitney U test (unadjusted p-values) with a prevalence of >= 50% were considered for the analyses. The visualisations were performed using R and Python. The generated data was resolved using the QIIME2 and mapped to the SILVA database. Predictive functional analysis was done using PICRUST2 across the KEGG pathways.

## 3. RESULTS

### 3.1 Descriptive data of recruited participants

The study population characteristics are mentioned in Table 1. Participants in the healthy and SAM groups were age and gender matched to improve the comparison between both the groups. Median age[months(IQR)] in healthy was [31.73(22.46, 40.23);n=72] and in SAM was [27.59(21.30,37.49);n=57). Gender distribution was balanced in both groups. The participants belonged to a similar economic background, irrespective of their current nutritional status.

### 3.2 Sociodemographic variables in recruited participants

In the univariate analysis, mother’s BMI, child’s birth weight, birth order, mode of delivery, and current breastfeeding status were each independently associated with SAM (Table 4). Specifically, for every 1 kg/m² increase in mother’s BMI, the odds of the child developing SAM decreased by 8% [Adjusted Odds Ratio (AOR): 0.92; 95% Confidence Interval (CI): 0.85–0.99]. Children with a birth weight of 2.5 kg or more had approximately 85% lower odds of developing SAM compared to those with lower birth weights [AOR: 0.15; 95% CI: 0.06–0.36]. In contrast, children who were third-born or later had about 4 times higher odds of developing SAM compared to firstborn children [AOR: 4.05; 95% CI: 1.33-12.30]. Additionally, children who were currently breastfeeding had around 3 times higher odds of SAM [AOR: 3.22; 95% CI: 1.21–8.61].

### 3.3 Dietary intake

Dietary data from 65 healthy and 51 SAM children (Tables 2 and 3) revealed significantly lower energy, carbohydrate, and protein intake in SAM children compared to healthy peers, with no difference in fat intake, AMDR, or dietary diversity. SAM children showed higher probabilities of inadequacy for multiple micronutrients, particularly thiamine, iron, zinc, and pyridoxine. Complementary feeding typically began between 5–7 months, predominantly with rice-based foods, and most SAM children were still breastfed at recruitment.

### 3.4 Gut Microbiota Diversity

Alpha diversity analysis (Figure 1A–C) revealed no significant differences between groups based on the Shannon (P=0.549) and Simpson (P=0.859) indices, indicating comparable richness and evenness of microbial communities. However, the Chao1 index, which is more sensitive to rare taxa, showed a statistically significant reduction in the SAM group (P=0.009), suggesting a depletion of low-abundance bacterial genera. We further analysed the data by stratifying samples based on age and gender(Figure S1-S2 A-B). In the 0–3 years age group, the SAM group exhibited significantly reduced diversity as measured by the Chao1 index (*P* = 0.03), whereas no significant differences were observed in the 4–5 years age group. Similarly, a significant reduction in diversity was observed among female subjects with SAM compared to healthy females (*P* = 0.03); however, no significant differences were detected among male subjects. Similarly, beta diversity analysis did not reveal any distinct clustering between groups (Figure 1G), as assessed by PERMANOVA (P=0.296). Overall, these results indicate that only minor differences in microbial diversity were observed between the groups. Additionally, genus-level relative abundance analysis (Figure 1F) revealed that *Bifidobacterium* was relatively more abundant in the SAM group, while genera such as *Blautia* and *Faecalibacterium* were more enriched in the healthy group. Together, these findings suggest that while overall diversity differences between groups are modest, the SAM-associated gut microbiome is characterised by reduced taxonomic richness and a loss of potentially beneficial genera, pointing to a less mature and potentially dysbiotic microbial community

**Figure 1:**
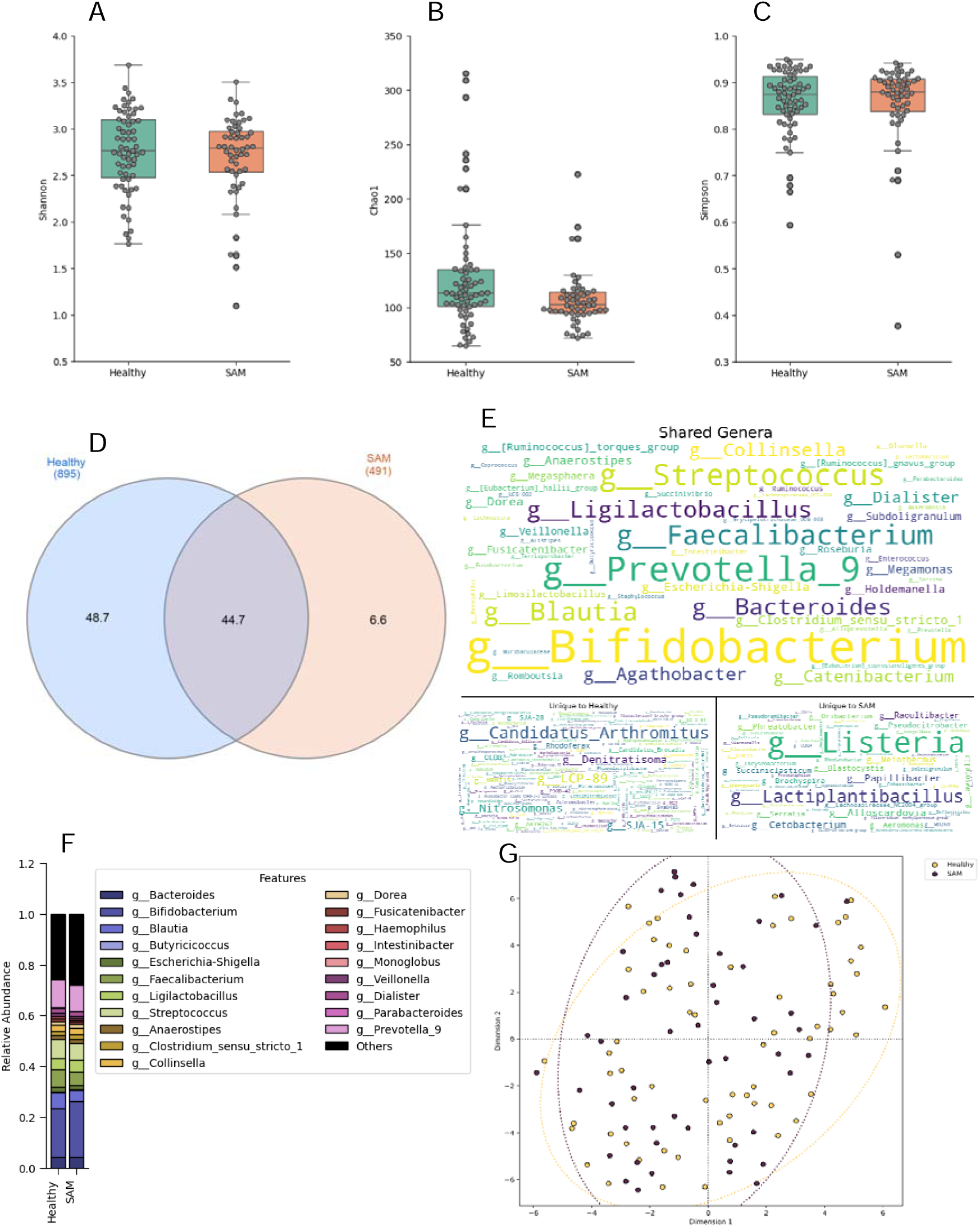
(A-G) Subtle Differences in Gut Bacteriome Diversity across the groups with indications of taxa loss in SAM. (A-C) The alpha diversity indices between Healthy and SAM group comparisons performed by Mann Whitney U test, (A) Shannon (P=0.549), (B) Chao1 (P<0.01), and (C) Simpson (P=0.859); (D-E) Shared and unique genera between SAM and Healthy, (D) Percentage wise, (E) Genera wise with larger fonts having higher abundance; (F) Relative abundance of top 20 genera having prevalence >75%; (G) Beta diversity analysis of bacteriome across groups, t-SNE used on calculated Bray-Curtis distances, followed by PERMANOVA test (P=0.296) the dashed ellipses define the outliers.

### 3.5 Differential abundance analyses

To further evaluate the compositional changes and shifts in the dynamics of intra-habitat microbial interactions, Lefse and Maslin3. The LefSe performed at a significance cut-off of 0.05 for Mann-whitney U test and a LDA score threshold of 3.0 identified an increase of *g_Limosilactobacillus, g_Enterococcus (f_Enterococcaceae), g_Succinivibrio (f_Succinivibrionaceae)* and g_*Staphylococcus (f_Staphylococcaceae; o_Staphylococcales)* in SAM. Whereas, *Blautia*, *Akkermansia* (*f_Akkermansiaceae*), *Ruminococcus gnavus group*, *Fusobacterium*, and the orders *Veillonellales-Selenomonadales* and *Fusobacteriales* were depleted in the gut of the same, while being enriched in the gut of children classified as healthy (Figure 2A). Stratified analyses based on age and gender revealed distinct patterns in microbial composition. In the **0–3 years** age group, *Limosilactobacillus* and *Staphylococcus* were significantly enriched in SAM, whereas *Megamonas*, *Fusobacterium*, and *Succinivibrio* were more abundant in healthy children. In the **4–5 years** age group, enrichment of *Muribaculaceae*, *Staphylococcus*, *Prevotella*, and *Limosilactobacillus* was observed in SAM, while *Romboutsia*, *Fusobacterium*, *Ruminococcus gnavus* group, *Erysipelotrichaceae UCG-003*, and *Akkermansia* were enriched in healthy children. Similarly, gender-based analyses showed that among female children, *Staphylococcus*, *Asteroleplasma*, *Turicibacter*, *Limosilactobacillus*, *Fusicanenibacter*, and the *Ruminococcus torques* group were enriched in SAM, whereas *Streptococcus*, *Escherichia-Shigella*, *Fusobacterium*, *Ruminococcus gnavus* group, *Akkermansia*, and *Erysipelotrichaceae UCG-003* were more abundant in healthy samples. Among **male children**, *Escherichia-Shigella* and *Limosilactobacillus* were enriched in SAM, while *Blautia*, *Fusicatenibacter*, *Ruminococcus*, *Parabacteroides*, and *CAG-352* were relatively more abundant in healthy counterparts. Further, using MaAsLin3 (Figure 2B), we modelled both the relative abundance (normalised using total sum scaling) and prevalence (logit-transformed binary outcome) of microbial features across the defined clinical categories. The concordant findings between LEfSe and MaAsLin3 were *g_Limosilactobacillus*, g_*Staphylococcus* and *Fusobacterium*. However, g_*Tyzzerella* was identified as enriched in healthy gut by MaAsLin3, which was not the case with LefSe. The varied abundances of the aforementioned microbial features reflect differences in gut environments and corresponding responses from the microbial community.

**Figure 2:**
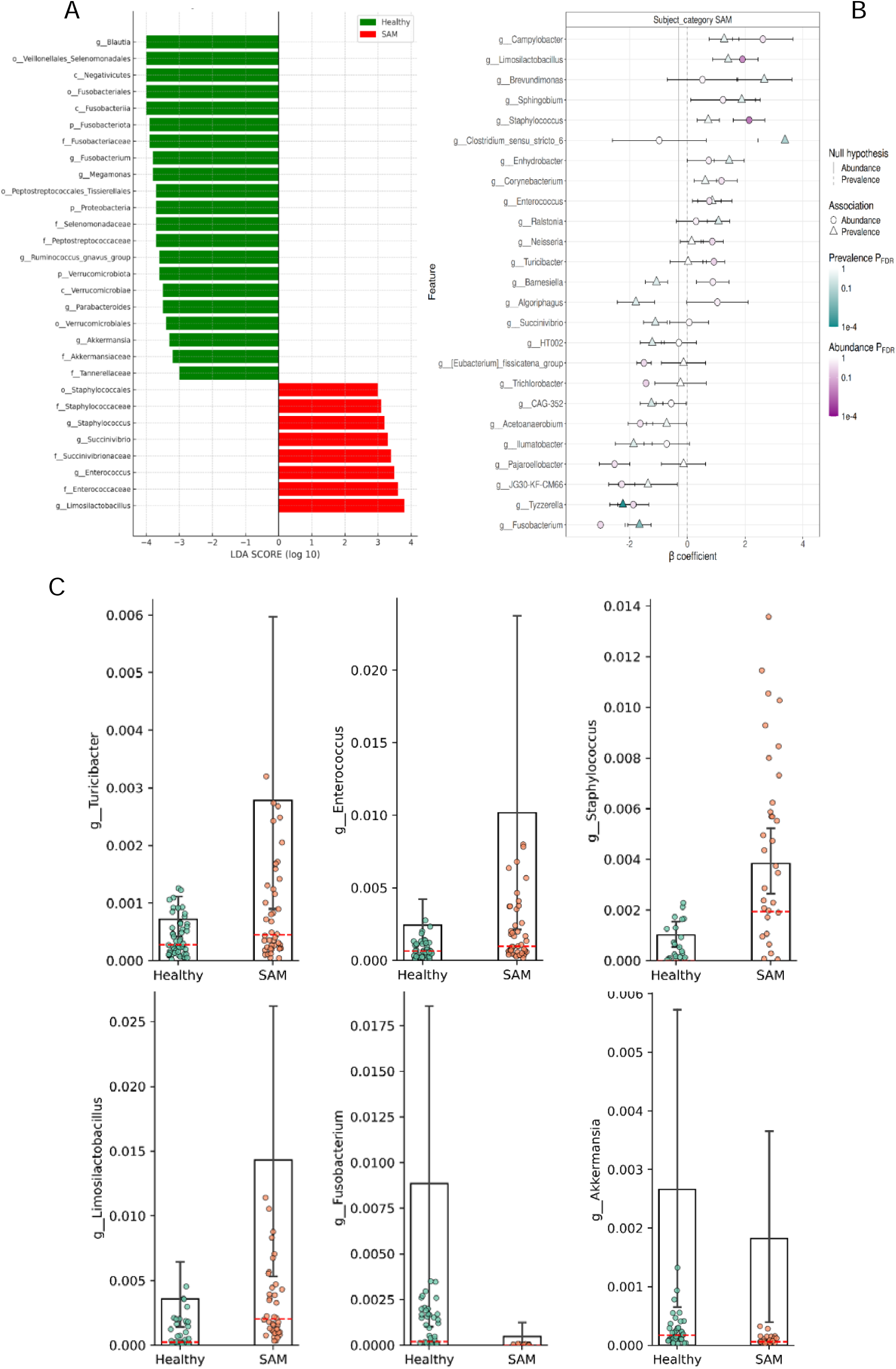
(A-B) Differential abundance analyses and gut microbial networks identify different interactions within intra-habitat ecosystems. (C) Represents the bacterial groups associated with SAM and study variables.

### 3.6 Inflammatory status in SAM and Healthy children

To assess the inflammatory status of the participants, we quantified three neutrophil-based inflammatory markers—myeloperoxidase (MPO), calprotectin (CAL), and neopterin (NEO)—along with secretory immunoglobulin A (sIgA) levels. Prior to statistical comparisons between children with Severe Acute Malnutrition (SAM) and healthy controls, outliers were removed from the healthy group using the interquartile range (IQR) method (1.5× the 25th and 75th percentiles), assuming that inflammation in apparently healthy children would remain within normal physiological limits. Group-wise comparisons were performed using the non-parametric Mann–Whitney U test. After outlier removal, the overall sample sizes for MPO, CAL, NEO, and sIgA analyses were as follows: Healthy (n = 37, 34, 36, and 40, respectively) and SAM (n = 41 for all). An increasing trend in inflammation was observed across all three markers in SAM children compared to healthy controls. Among these, MPO and CAL were significantly elevated in the SAM group (p = 0.026 and p = 0.014, respectively; Figure S7), while NEO levels were higher in SAM but did not reach statistical significance. In contrast, sIgA levels were lower in SAM children compared to healthy controls.

Gender-stratified analysis showed no significant differences among females; however, among males, MPO and CAL levels were significantly higher in the SAM group compared to healthy males (Figures S8 and S9). For MPO, CAL, and NEO, the male subgroup sample sizes were 18–20 per group, while for females they ranged between 18–21 per group. Age-based stratification revealed that the greatest inflammatory burden was observed in younger SAM children (0–3 years), who exhibited significantly higher MPO and CAL levels compared to age- and gender-matched healthy controls (Figure S10 and S11). For this age group, sample sizes ranged from 22–25 per group, while for the 4–5 year group, they ranged from 13–16 per group. Overall, these findings indicate heightened intestinal inflammation in SAM children, particularly in younger age groups and among males, with MPO and CAL serving as key markers of this elevated inflammatory state.

### 3.7 Correlation between Clinical, Diet, Sociodemographic variables, fecal inflammatory markers and Bacterial abundance

Associations between measured parameters and gut bacterial genera define different behaviours of genera in the changing gut environment.

We analysed the relationship between gut microbial genera and host parameters, including dietary, anthropometric measures, nutrient intake and inflammatory biomarkers using Spearman correlation(Figure 3A). Among the significantly associated genera, *Enterococcus, Staphylococcus, Limosilactobacillus* and *Turicibacter* are negatively associated with child growth metrics like **WHZ, WAZ and HAZ** scores, suggesting a possible role in undernutrition and growth faltering. Within these groups, the negative association between Enterococcus abundance and nutrient intake, particularly energy, protein, and fat, suggests that higher Enterococcus levels may be linked to reduced dietary intake or impaired nutrient absorption. This pattern could reflect a dysbiotic microbial environment that negatively influences host nutritional status or vice versa. On the contrary, genera *Ruminococcus gnavus group*, *Blautia, Akkermansia, Colidextribacter and Elisipelotrichaeceae_UGC_003* are positively associated with weight and other growth measures like **WHZ, HAZ and WAZ** scores.

**Figure 3:**
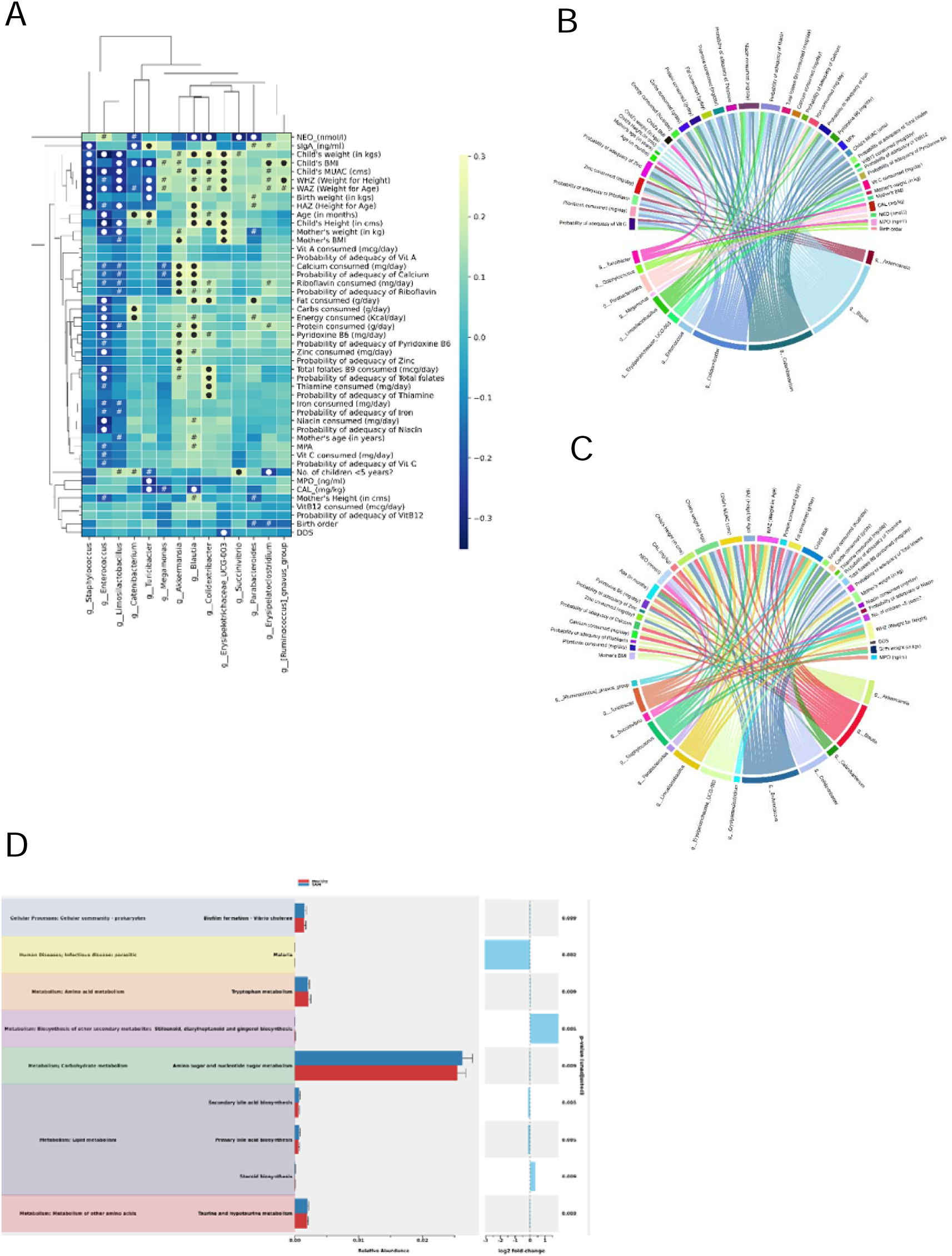
(A-D) Differential abundance analyses and gut microbial networks identify different interactions within intra-habitat ecosystems. (A-C) Spearman Correlation analysis was performed on features with prevalence > 50% and a significant difference in the Mann-Whitney U test between the Healthy and SAM groups; (A) The color demonstrates the strength and direction of the associations, while the mask ‘#’ indicates p-value < 0.1 obtained in correlation analyses and ‘●’ indicates p-value < 0.05. All other associations are non-significant; (B) Non-directional weighted significant associations at p-value <= 0.05 by sub-setting dataset to SAM group; (C) Non-directional weighted significant associations at p-value <= 0.05 by sub-setting dataset to Healthy group. (D) Predictive functional analysis of gut bacterial communities from SAM and healthy children was conducted using PICRUSt2

In relation to inflammation, *Blautia* and *Colidextribacter* were inversely correlated with fecal neopterin and calprotectin levels, suggesting their protective, anti-inflammatory potential. Notably no bacterial taxa were found to be positively associated with these inflammatory markers.

*Turicibacter* was found to be negatively associated with WHZ and WAZ, which are critical markers for acute malnutrition. It was also found to be positively associated with fecal secretory IgA (sIgA) levels. It’s significant enrichment in SAM children (p=0.03) suggests a possible role in pathophysiology of undernutrition. This association warrants further investigation.to clarify the underlying mechanisms.

Enterococcus, a facultative anaerobe, was enriched in the gut microbiota of children with SAM and relatively depleted in the healthy group. Its abundance showed a negative association with growth metrics of the child and the maternal weight. Similarly, it is inversely associated with energy, protein, and fat intake in children, suggesting its possible role in undernutrition. Predictive functional analysis of gut bacterial communities from SAM and healthy children was conducted using PICRUSt. Functional prediction identifies liver-related metabolism to be altered, indicating a further role of the gut-liver axis. Figure 3D presents both Level 1 and Level 2 KEGG pathway data, highlighting the functional pathways with significant differences in their relative abundances between SAM and healthy children (p < 0.05). Level 2 pathways such as Biofilm formation - *Vibrio cholerae*; infectious diseases-malaria; amino sugar and nucleotide sugar metabolism; primary and secondary bile acid syntheses; taurine and hypotaurine metabolism were higher in abundances in SAM group, whereas, pathways such as tryptophan metabolism; stilbenoid, diarylheptanoid and gingerol biosynthesis; steroid biosynthesis were higher in abundances in healthy group. However, the aforementioned observations must be handled with caution due to the predictive nature of the analysis.

## 4. Discussion

Our study highlights the complex interplay between diet, socioeconomic factors, inflammation, and bacterial dynamics in the pathophysiology of undernutrition. Socioeconomic determinants influencing undernutrition are diverse, including maternal education and health status, family size, birth order and birth weight of the child.(19–21). Consistent with these parameters, our findings reveal that maternal BMI is negatively associated with risk of SAM, indicating that poor nutritional status may predispose children to undernutrition (22). Additionally, higher Birth order (3 or beyond) shows a positive association with SAM (21), consistent with previous studies from India and other parts of South Asia (24–26). This elevated risk among later-born children is often linked to short birth intervals, leading to inadequate maternal recovery, nutrient depletion, and limited household resources distributed across multiple dependents. These factors collectively heighten vulnerability to malnutrition, emphasizing the importance of maternal nutrition and family planning interventions in reducing SAM, particularly in larger families. Our data also revealed that current breastfeeding status was positively associated with SAM, possibly reflecting compensatory feeding behaviour by mothers. This observation warrants further exploration into breastfeeding practices, maternal nutrition, and feeding adequacy in this population.

Dietary analysis demonstrated that children with SAM consumed significantly lower amounts of carbohydrates, protein, and overall energy, compared to healthy controls, indicating inadequate macronutrient intake. Despite comparable dietary diversity between groups, both exhibited insufficiency across multiple micronutrients, as indicated by the mean probability of adequacy (MPA) scores. The findings were consistent with the previous reports, reporting widespread micronutrient deficiency among SAM children(12,27). Dietary diversity is known to be directly proportional to microbial diversity and abundance(28). Despite similar dietary diversity between both the groups, we have observed that SAM is associated with altered gut microbial diversity (Figure 1). This can likely be driven by nutrient deficiency, intestinal barrier dysfunction, environmental factors, impaired gut health, or immune system impairment (12,15,29). These insights suggest the need to view dietary diversity alongside nutrient adequacy and gut functional health when addressing undernutrition.

Comparison of the gut bacterial diversity between SAM and healthy children showed significantly reduced microbial richness in the SAM group as measured by Chao1 index. This aligns with the previous findings that reported decreased microbial diversity in SAM, indicative of delayed or impaired gut ecosystem development. However, it is important to note that few studies have reported no significant difference in microbial diversity between these groups indicating variability in findings possibly due to differences in study populations, or methodologies (12,15,30). We found that the unique bacterial taxa to SAM accounted for only 6.6% of the total microbiota, suggesting that although overall diversity declines, the core microbial structure remains largely conserved, with potential loss of specific beneficial taxa critical for gut functions.

Maternal factors, including diet, plays a crucial role shaping the infant gut-microbial community development further influencing susceptibility to infections and disease severity (31). A recent study from West Africa reported a higher abundance of *Listeria monocytogenes* in breast milk of mothers with SAM-affected children (32). Children with SAM are therefore at a relatively higher risk of L. monocytogenes infiltration, possibly due to reduced diversity and depletion of Lactobacillus species, which normally exerts inhibitory effects against Listeria growth (3,14). In accordance to this finding, though inconsistent across all cases, our dataset has revealed the presence of *Listeria* in a few SAM samples. This highlights the potential role of pathogenic bacteria in the compromised gut environment of SAM children and demands further investigation through future studies.

Further analysis through LefSe has shown an enrichment of *Staphylococcus, Enterococcus* and *Limosilactobacillus* in SAM group. *Staphylococcus* is noted for its pathogenic traits by reducing butyrate production, a key short-chain fatty acid (SCFA) essential for maintaining gut health, regulating inflammation, and supporting nutrient absorption. Its increased abundance in SAM children is indicative of its contribute to intestinal inflammation and malabsorption, exacerbating malnutrition. (33,34). Similarly, *Enterococcus*, a subdominant commensal, can behave as an opportunistic pathogen under nutrient stress or antibiotic exposure, disrupting microbial balance and inducing inflammation (14). Its proliferation can result in the disruption of gut homeostasis, leading to production of antimicrobial peptides and biofilms, making it resistant to the host defenses. (35–37). We have also observed a negative correlation of Enterococcus abundance with dietary intake and growth matrices. Together these findings suggest that overrepresentation of these opportunistic groups make them act as a key disruptor of gut function in SAM, intensifying inflammation, malabsorption, and overall nutritional decline.

In addition to these pathogenic signatures, we found increased abundance of *Turicibacter* in the SAM group. These genus thrives in serotonin-rich gut environments and carries the transporter protein CUW_0748, homologous to the host serotonin reuptake receptor (SERT), facilitating serotonin uptake (38–40). This shared affinity for serotonin between *Turicibacter* and host enterocytes may create competition that alters gut motility and transit time, potentially reducing nutrient absorption efficiency (41). Our findings indicate that increased *Turicibacter* levels can possibly impact gut motility reflected by an increased frequency of defecation per day in SAM children. Literature findings support this mechanism, showing that an increased *Turicibacter* abundance leads to accelerated intestinal transit time (42).

Inflammatory marker assessment performed in a subset of samples has shown a clear trend of increased inflammation among SAM children compared to healthy controls, in accordance to the previous literature findings (43,44). Increased inflammation is closely linked to increased permeability, which can further impair nutrient absorption (45,46). Gender based analysis has shown that male children exhibited higher levels of inflammation than their female counterparts. Latest data from the MAL-ED study also reported increased intestinal permeability in undernourished children, being more pronounced among males than females(47). In addition, we have found reduced fecal sIgA levels in the SAM group. Furthermore, a gender-based comparisons have revealed a significant difference in the male groups. Altogether, these findings suggest that male children may face an elevated risk of malnutrition due to both, heightened inflammation and decreased sIgA levels. Supporting this observation, a recent meta-analysis confirmed that males have higher odds of experiencing undernutrition compared to females(48).

Functional prediction revealed diminished tryptophan(trp) metabolism in SAM consistent with previous reports (49,50). Altered bacterial tryptophan metabolism is known to disrupt gut barrier integrity and CD4+ T-cell differentiation (51). Altered tryptophan catabolism affects immune modulation and gut integrity by disrupting CD4+ T-cell differentiation and epithelial barrier stability. Furthermore, metabolic pathway analysis indicated disturbed lipid metabolism, particularly increased primary and secondary bile acid biosynthesis, an indicative of bile acid malabsorption and impaired reuptake (52). Elevated bile acid levels have been associated with dysbiosis in SAM (53). Similarly, *Turicibacter* enrichment has been found to be associated with bile acid alteration and lipid imbalance (54). These reportings provide mechanistic insights into how gut dysbiosis exacerbates the physiological effects of undernutrition.

Our study outcomes highlight how factors such as socio-economic determinants, dietary quality, and gut microbial disruptions tend to influence vulnerability and outcomes in these nutritionally compromised individuals. The findings reinforce that maternal nutrition, education, and family planning are critical for prevention, while the roles of gut dysbiosis, impaired barrier function, and inflammation act as mediators of nutritional deficits and recovery trajectories. Addressing undernutrition effectively will require integrated strategies that encompass social, nutritional, and microbiome-targeted interventions to break the cycle of malnutrition and promote healthier growth in populations at risk.

## Supporting information

Supplementary tables and CONSORT

Questionnaire for participant

## Data Availability

All data produced in the present study are available upon reasonable request to the corresponding author.

