## Supplementary tables and CONSORT for "Gut Microbial Dysbiosis and Elevated Mucosal Inflammation in Severe Acute Malnutrition: A Case-Control Study from India"

| **sVariables** | **Healthy**  **(n= 72)** | **SAM**  **(n= 57)** | **p-value** |
| --- | --- | --- | --- |
| **Child Age (in months), median (IQR)** | 31.73  (22.46, 40.23) | 27.59  (21.30, 37.49) | 0.505 |
| **Gender, n (%)** |  | | |
| Female | 36 (50.0) | 29 (50.9) | 0.921 |
| Male | 36 (50.0) | 28 (49.1) |  |
| **Child’s height (in cm), median (IQR)** | 87.4  (81.4, 94.9) | 82.5  (76.0, 89.0) | 0.022 |
| **Child’s weight (in kg), median (IQR)** | 12.0  (10.5, 13.4) | 8.2  (7.3, 9.2) | <0.001 |
| **Child’s MUAC (in cm), median (IQR)** | 14.6  (14.0, 15.4) | 12.1  (11.5, 12.6) | <0.001 |
| **Child’s BMI (kg/m2), median (IQR)** | 15.24  (14.52, 16.28) | 12.08  (11.81, 12.47) | <0.001 |

**Table. 1 Descriptive characteristics of study participants**

**Table. 2 Nutrient consumption pattern among study participants**

| **Variables** | **Healthy**  **(n= 65)** | **SAM**  **(n= 51)** | **p-value** |
| --- | --- | --- | --- |
| **Energy consumption (in Kcal), median (IQR)** | 1254.49 (923.76, 1582.74) | 1072.31 (806.32,  1503.03) | **0.037** |
| **Energy gap (in Kcal), median (IQR)** | 77.58  (-229.55, 392.20) | -190.31  (-361.36, 194.90) | **0.015** |
| **Carbohydrate consumption (in grams), median (IQR)** | 177.58  (128.02, 213.11) | 137.81 (102.78, 191.33) | **0.022** |
| **Percent Energy from Carbs, median (IQR)** | 55.88  (49.13, 60.92) | 53.39  (47.59, 63.99) | 0.666 |
| **Protein consumption (in grams), median (IQR)** | 33.76  (24.95, 40.07) | 26.85  (17.67, 36.47) | **0.011** |
| **Percent Energy from Protein, median (IQR)** | 10.66  (9.39, 11.79) | 10.00  (8.43, 11.24) | 0.120 |
| **Fat consumption (in grams), median (IQR)** | 36.88  (25.89, 55.09) | 32.96  (21.56, 53.25) | 0.396 |
| **Percent Energy from Fat, median (IQR)** | 28.25  (21.82, 33.47) | 30.51  (22.41, 37.08) | 0.157 |
| **MPA, median (IQR)** | 0.0948  (0.0141, 0.2374) | 0.0795 (0.0005, 0.1819) | **0.032** |
| **DDS, median (IQR)** | 5 (4, 5) | 5 (4, 6) | 0.108 |

**Table.3 Dietary determinants of study participants**

| **Variables** | **Healthy**  **(n= 65)** | **SAM**  **(n= 51)** | **p-value** |
| --- | --- | --- | --- |
| **Macronutrient Distribution Range and Macronutrient Adequacy** | | | |
| **AMDR carbohydrate status, n (%)** | | | |
| Satisfied | 45 (69.2) | 30 (58.8) | 0.244 |
| **AMDR Protein status, n (%)** | | | |
| Satisfied | 65 (100.0) | 51 (100.0) | **-** |
| **AMDR Fat status, n (%)** | | | |
| Satisfied | 22 (33.9) | 24 (47.1) | 0.149 |
| **Probability of Inadequacy of Micronutrients, median (IQR)** | | | |
| Vitamin A | 0.983(0.941) | 0.991(0.211) | 0.162 |
| Vitamin C | 0.984(0.991) | 1.00(1.00) | 0.532 |
| Thiamine | 0.999(0.563) | 1.00(0.025) | **0.047** |
| Riboflavin | 1.00(1.50 x 10^-4^) | 1.00(2.31 x 10^-7^) | 0.071 |
| Niacin | 0.999(0.195) | 0.999(0.028) | 0.110 |
| Total Folates | 1.00(6.69 x 10^-4^) | 1.00(1.15 x 10^-4^) | 0.230 |
| Vitamin B12 | 1.00(3.22 x 10^-15^) | 1.00(2.62 x 10^-11^) | 0.620 |
| Calcium | 1.00(0.089) | 1.00(3.22 x 10^-4^) | 0.286 |
| Iron | 1.00(0.035) | 1.00(0.002) | **0.024** |
| Zinc | 1.00(1.08 x 10^-11^) | 1.00(9.88 x 10^-15^) | **0.020** |
| Pyridoxine | 1.00(4.62 x 10^-4^) | 1.00(6.40 x 10^-8^) | **0.021** |
| **Dietary Diversity Score, n (%)** | | | |
| Good (≥ 4) | 60 (92.3) | 48 (94.1) | 1.000 |
| Poor | 5 (7.7) | 3 (5.9) |  |
| **Infant feeding and child dietary practices** | | | |
| **Age of introduction of complementary feeding, n (%)** | | | |
| < 5 months | 4 (5.6) | 1 (1.7) | 0.137 |
| 5 to 7 months | 62 (87.3) | 46 (80.7) |  |
| >7 months | 5 (7.0) | 10 (17.5) |  |
| **Major complimentary food, n (%)** | | | |
| Rice based | 53 (74.6) | 45 (78.9) | 0.947 |
| Rice based/millet based | 1 (1.4) | 0 (0.0) |  |
| Rice based/Wheat based | 13 (18.3) | 10 (17.5) |  |
| Wheat based | 3 (4.2) | 1 (1.7) |  |
| Others | 1 (1.4) | 1 (1.7) |  |
| **Still breastfeed, n (%)** | | | |
|  | 17 (23.9) | 31 (54.4) | **<0.001** |

**Table.4 Univariable analysis of sociodemographic and dietary factors**

| **Variables** | **OR (95% CI)** | **p-value** |
| --- | --- | --- |
| **Mother’s age (years)** | 1.04 (0.92, 1.17) | 0.531 |
| **Mother’s BMI (kg/m2)** | 0.92 (0.85, 0.99) | **0.034** |
| **No. of children mother having** | | |
| 0 | Ref | - |
| ≤2 | 1.32 (0.12, 15.18) | 0.824 |
| >2 | 2.29 (0.19, 28.0) | 0.518 |
| **Children less than 5 years** | | |
| 1 | Ref | - |
| 2 | 0.69 (0.31, 1.51) | 0.347 |
| 3 | 0.25 (0.03, 2.42) | 0.231 |
| **Birth Order** | | |
| 1 | Ref | - |
| 2 | 1.30 (0.60, 2.82) | 0.503 |
| 3 | 4.05 (1.33, 12.30) | **0.014** |
| **Mode of delivery** | | |
| Vaginal/Episiotomy | Ref | **0.006** |
| Caesarean Section | 0.35 (0.17, 0.74) |  |
| **Birth weight (Kg)** | | |
| $<$2.5 kg | Ref | **<0.001** |
| $\geq$2.5 kg | 0.15 (0.06, 0.36) |  |
| **Still breastfeed** | | |
| No | Ref | **0.001** |
| Yes | 3.79 (1.78, 8.05) |  |
| **Energy consumption (in Kcal)** | | |
| Per 10 Kcal | 0.99 (0.983, 0.999) | **0.034** |
| Per 100 Kcal | 0.91 (0.84, 0.99) | **0.034** |
| **AMDR carbohydrate status** | | |
| Satisfied | Ref | 0.246 |
| Not Satisfied | 1.56 (0.73, 3.39) |  |
| **AMDR Protein status** | | |
| Satisfied | Can not be calculated | **-** |
| Not Satisfied |  |  |
| **AMDR Fat status** | | |
| Satisfied | Ref | 0.150 |
| Not Satisfied | 0.58 (0.27, 1.22) |  |
| **MPA Status** | | |
| ≥ 0.5 | Ref | **0.044** |
| < 0.5 | 2.57 (1.02, 6.41) |  |
| **DDS status** | | |
| Good | Ref | 0.703 |
| Poor | 0.75 (0.17, 3.30) |  |

**S3 Equation for calculation of raw food amount from consumed cooked food.**

*Quantity of raw food ingredient consumed = (Quantity of raw food ingredient cooked/Total quantity of cooked food) * Quantity of cooked food consumed.*

**S4 CONSORT DIAGRAM OF THE STUDY**

**CONSORT Flowchart of the study**

**Anganwadi centers screened-70**

Children assessed for eligibility (n=341)

**Analysis**

Recruited (n= 129)

**Diet data collected (n=52)**

**Fecal sample collection & Analysis (n=63)**

**Fecal sample collection & Analysis (n=52)**

Excluded (n=212)

- Not meeting inclusion criteria

- Consumed Antibiotic/probiotic in last 4 weeks

**Diet data collected (n=65)**

**SAM (n= 57)
Assessed for clinical and sociodemographic information**

**Healthy (n=72)
Assessed for clinical and sociodemographic information**
