## Supplementary figures for "Gut Microbial Dysbiosis and Elevated Mucosal Inflammation in Severe Acute Malnutrition: A Case-Control Study from India"

### Slide 1
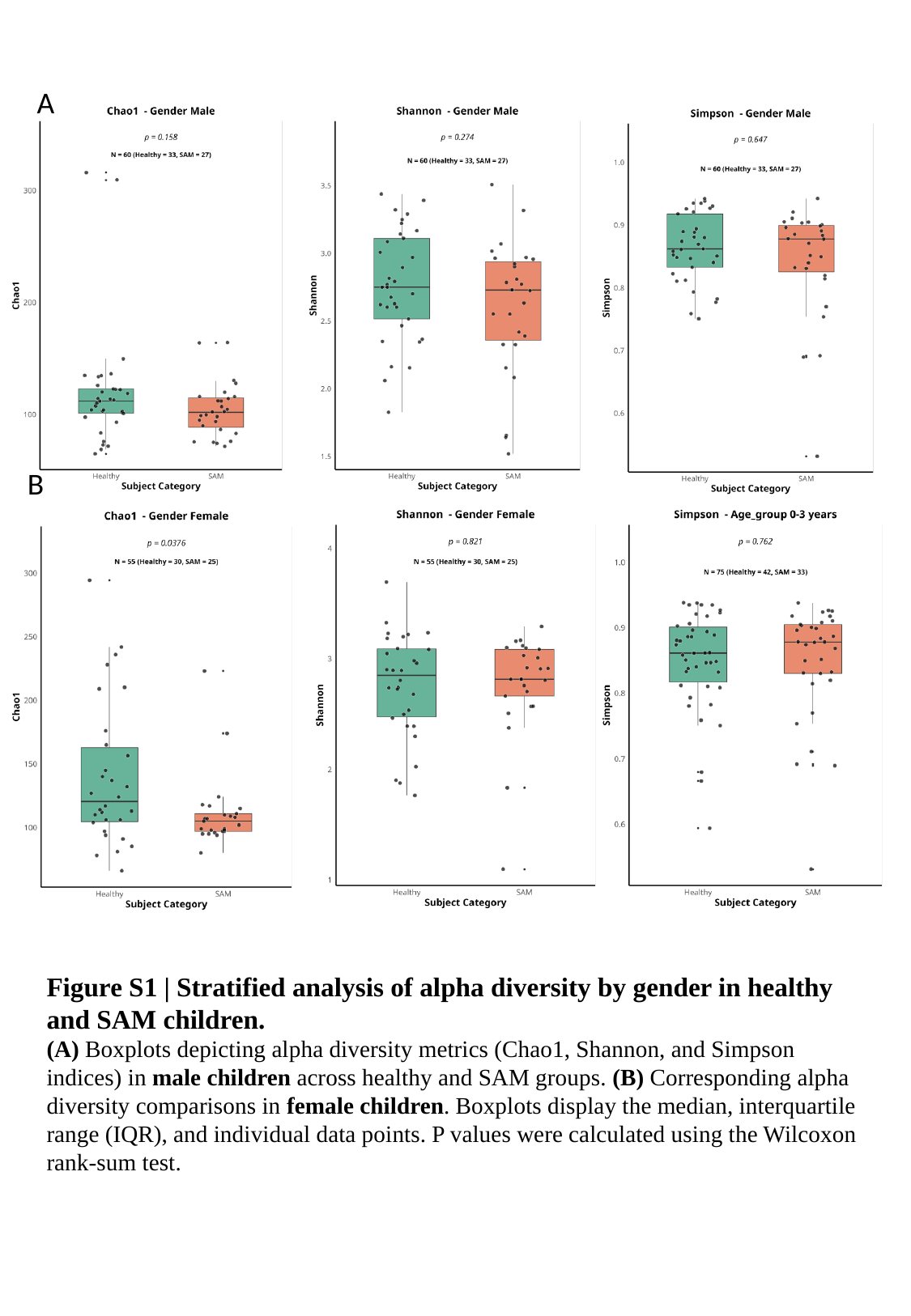

A
B
Figure S1 | Stratified analysis of alpha diversity by gender in healthy and SAM children.
(A) Boxplots depicting alpha diversity metrics (Chao1, Shannon, and Simpson indices) in male children across healthy and SAM groups. (B) Corresponding alpha diversity comparisons in female children. Boxplots display the median, interquartile range (IQR), and individual data points. P values were calculated using the Wilcoxon rank-sum test.

### Slide 2
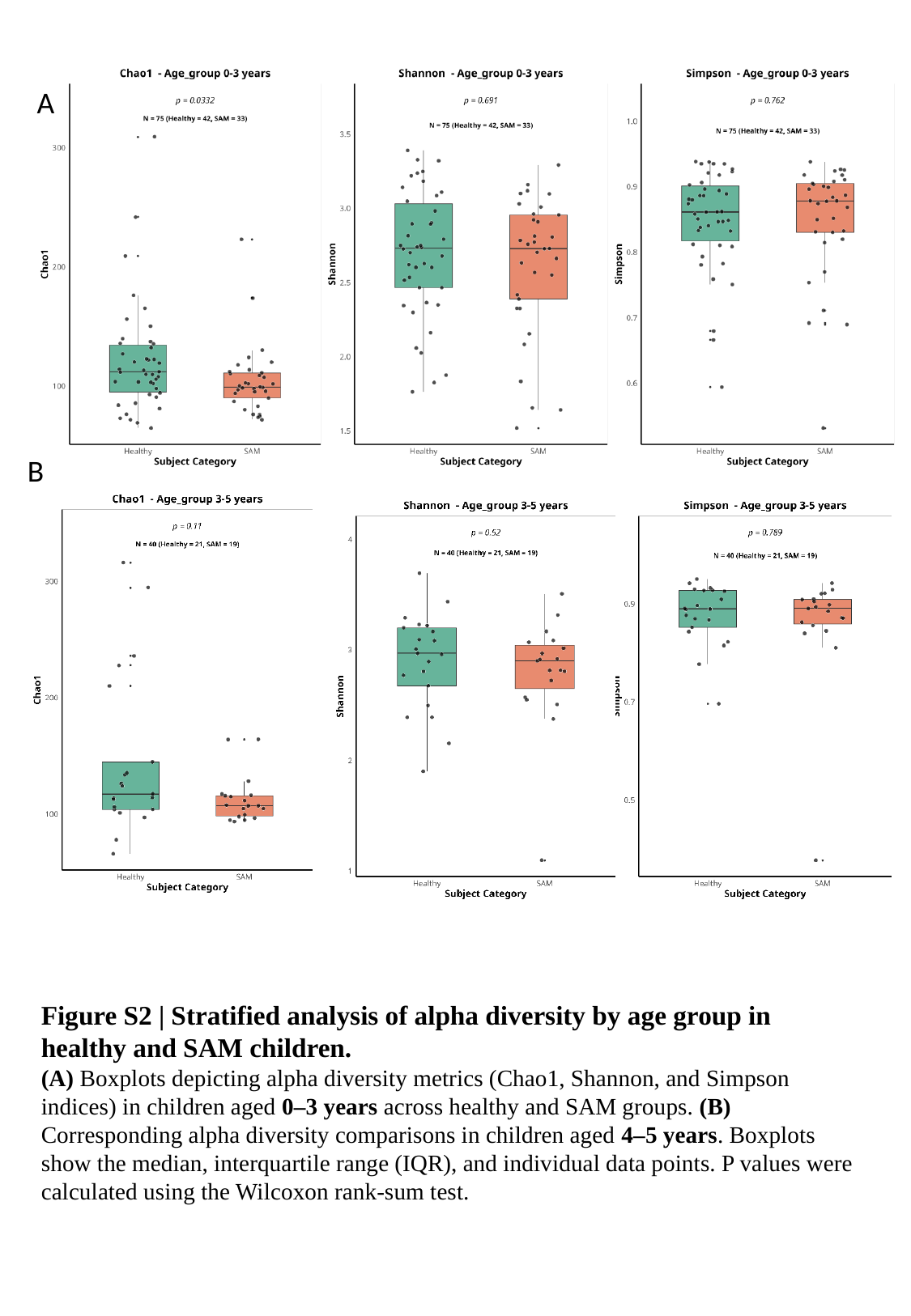

A
B
Figure S2 | Stratified analysis of alpha diversity by age group in healthy and SAM children.
(A) Boxplots depicting alpha diversity metrics (Chao1, Shannon, and Simpson indices) in children aged 0–3 years across healthy and SAM groups. (B) Corresponding alpha diversity comparisons in children aged 4–5 years. Boxplots show the median, interquartile range (IQR), and individual data points. P values were calculated using the Wilcoxon rank-sum test.

### Slide 3
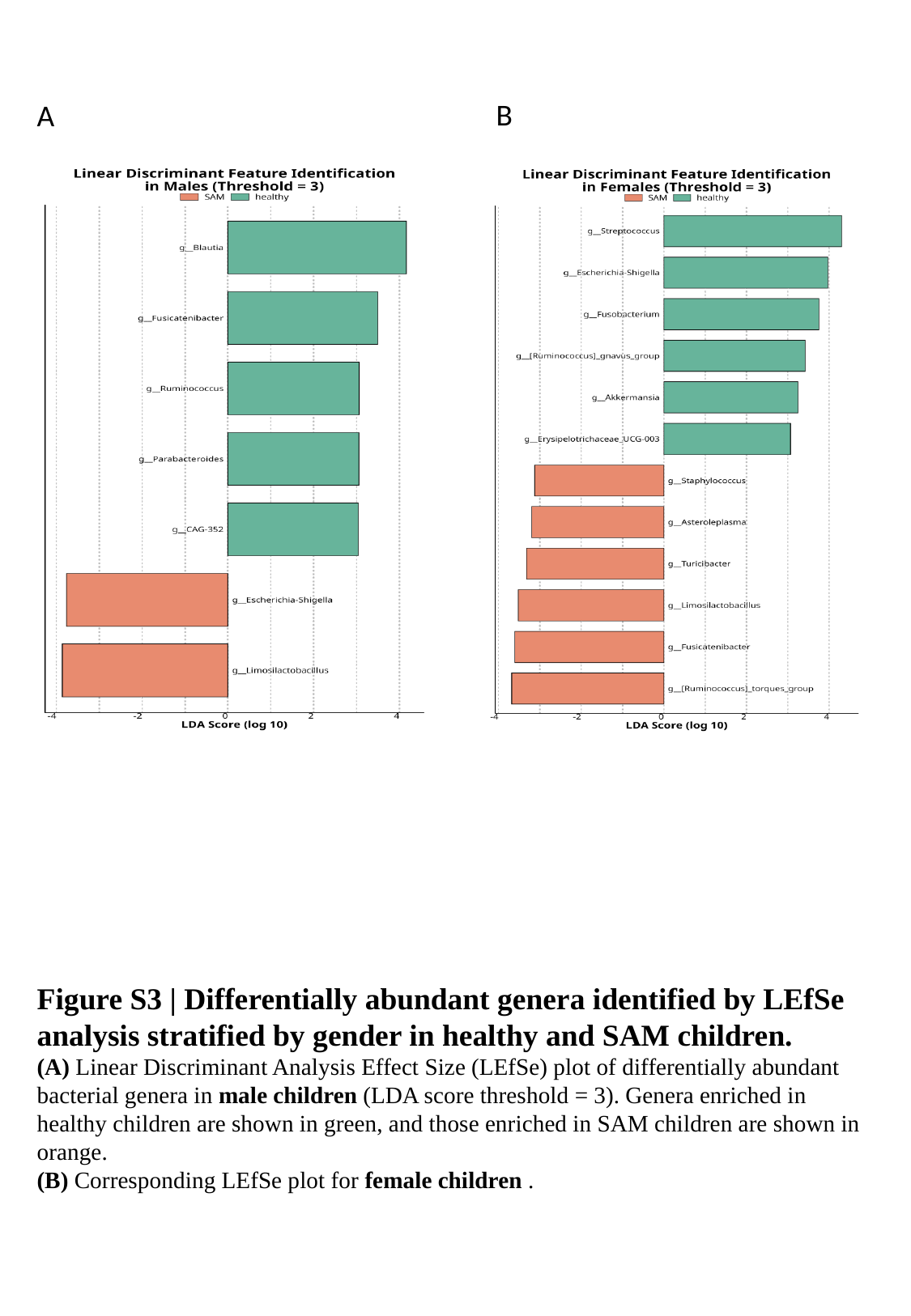

B
A
Figure S3 | Differentially abundant genera identified by LEfSe analysis stratified by gender in healthy and SAM children.
(A) Linear Discriminant Analysis Effect Size (LEfSe) plot of differentially abundant bacterial genera in male children (LDA score threshold = 3). Genera enriched in healthy children are shown in green, and those enriched in SAM children are shown in orange.
(B) Corresponding LEfSe plot for female children .

### Slide 4
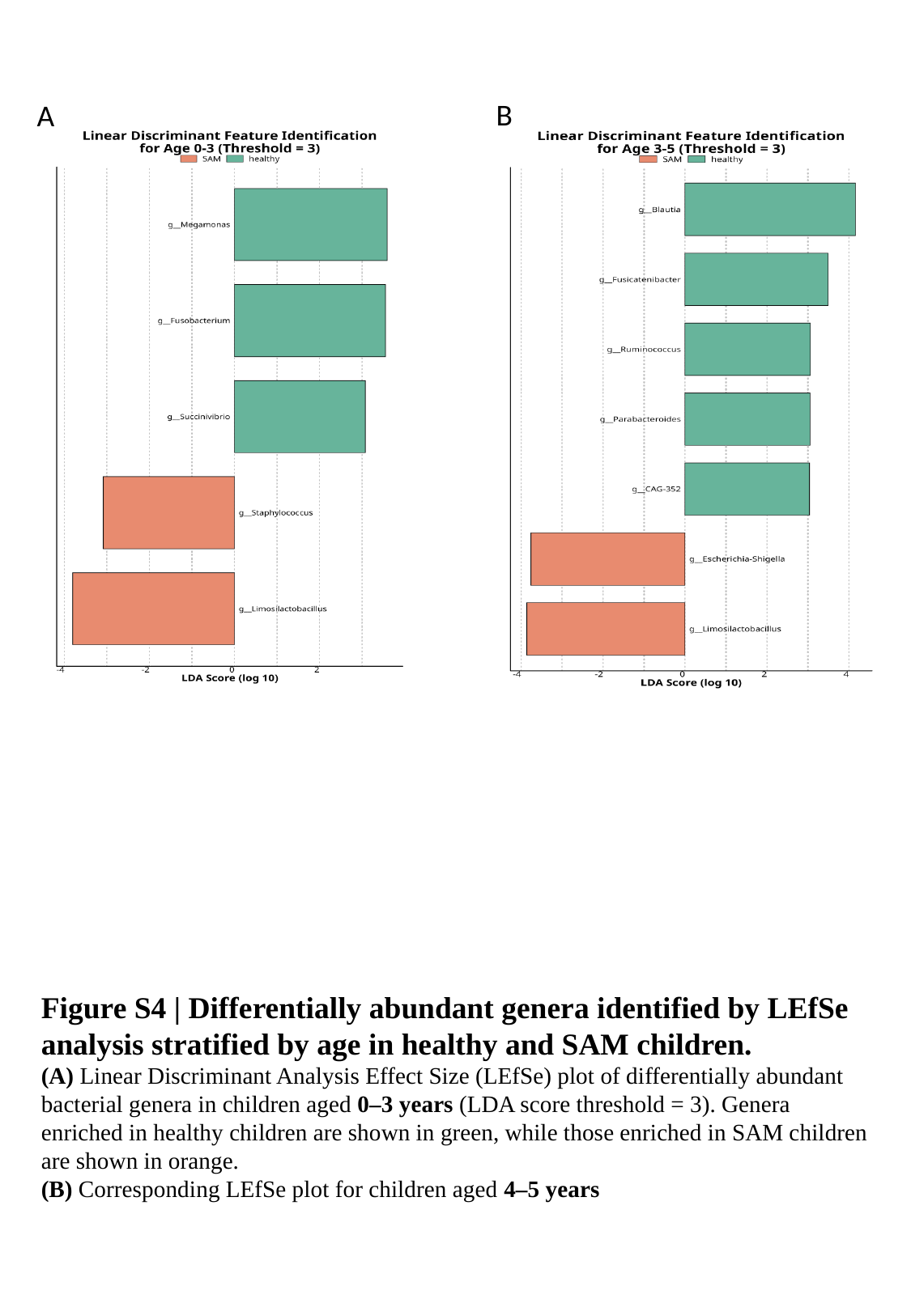

B
A
Figure S4 | Differentially abundant genera identified by LEfSe analysis stratified by age in healthy and SAM children.
(A) Linear Discriminant Analysis Effect Size (LEfSe) plot of differentially abundant bacterial genera in children aged 0–3 years (LDA score threshold = 3). Genera enriched in healthy children are shown in green, while those enriched in SAM children are shown in orange.
(B) Corresponding LEfSe plot for children aged 4–5 years

### Slide 5
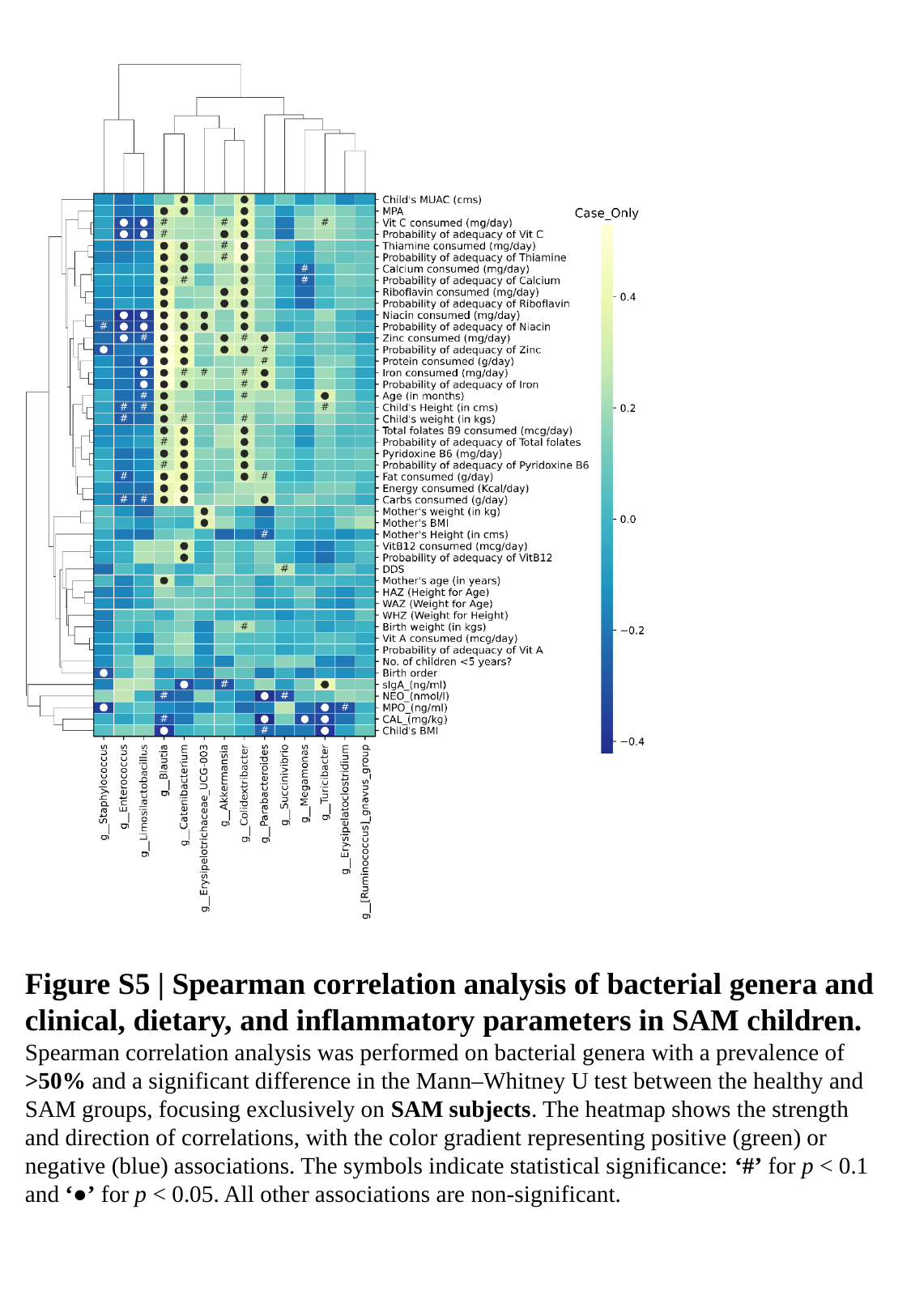

Figure S5 | Spearman correlation analysis of bacterial genera and clinical, dietary, and inflammatory parameters in SAM children.
Spearman correlation analysis was performed on bacterial genera with a prevalence of >50% and a significant difference in the Mann–Whitney U test between the healthy and SAM groups, focusing exclusively on SAM subjects. The heatmap shows the strength and direction of correlations, with the color gradient representing positive (green) or negative (blue) associations. The symbols indicate statistical significance: ‘#’ for p < 0.1 and ‘●’ for p < 0.05. All other associations are non-significant.

### Slide 6
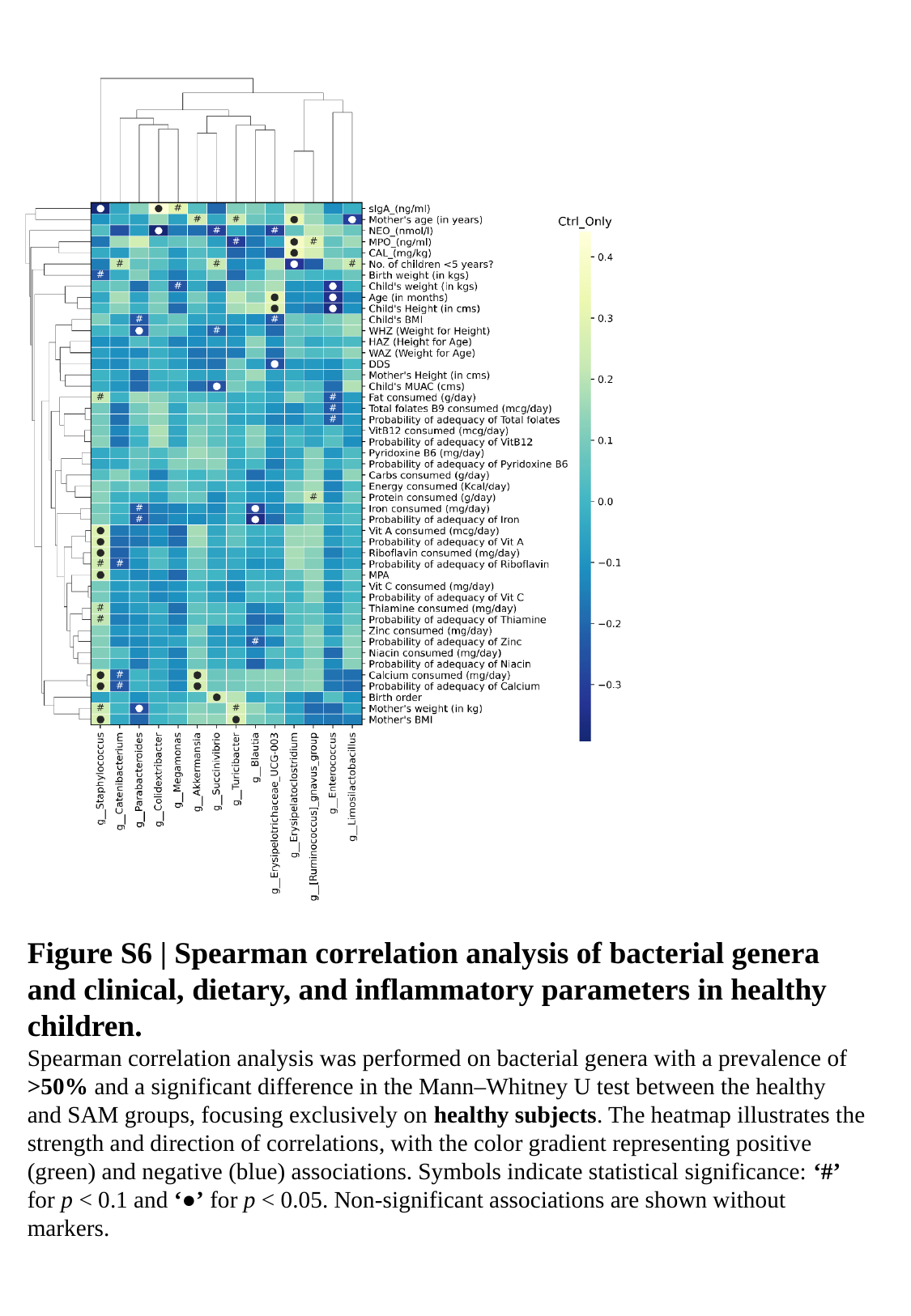

Figure S6 | Spearman correlation analysis of bacterial genera and clinical, dietary, and inflammatory parameters in healthy children.
Spearman correlation analysis was performed on bacterial genera with a prevalence of >50% and a significant difference in the Mann–Whitney U test between the healthy and SAM groups, focusing exclusively on healthy subjects. The heatmap illustrates the strength and direction of correlations, with the color gradient representing positive (green) and negative (blue) associations. Symbols indicate statistical significance: ‘#’ for p < 0.1 and ‘●’ for p < 0.05. Non-significant associations are shown without markers.

### Slide 7
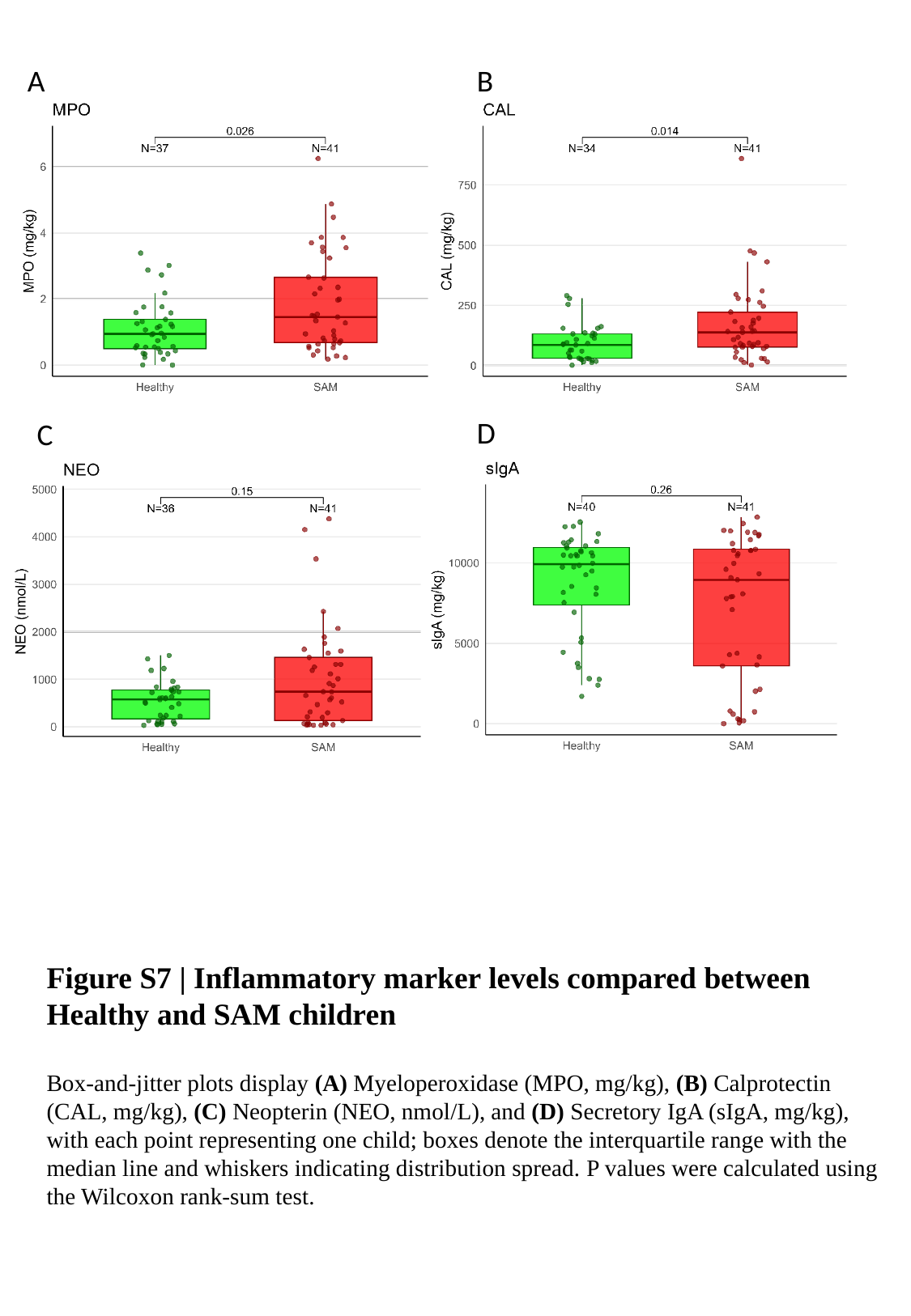

A
B
D
C
Figure S7 | Inflammatory marker levels compared between Healthy and SAM children
Box‑and‑jitter plots display (A) Myeloperoxidase (MPO, mg/kg), (B) Calprotectin (CAL, mg/kg), (C) Neopterin (NEO, nmol/L), and (D) Secretory IgA (sIgA, mg/kg), with each point representing one child; boxes denote the interquartile range with the median line and whiskers indicating distribution spread. P values were calculated using the Wilcoxon rank-sum test.

### Slide 8
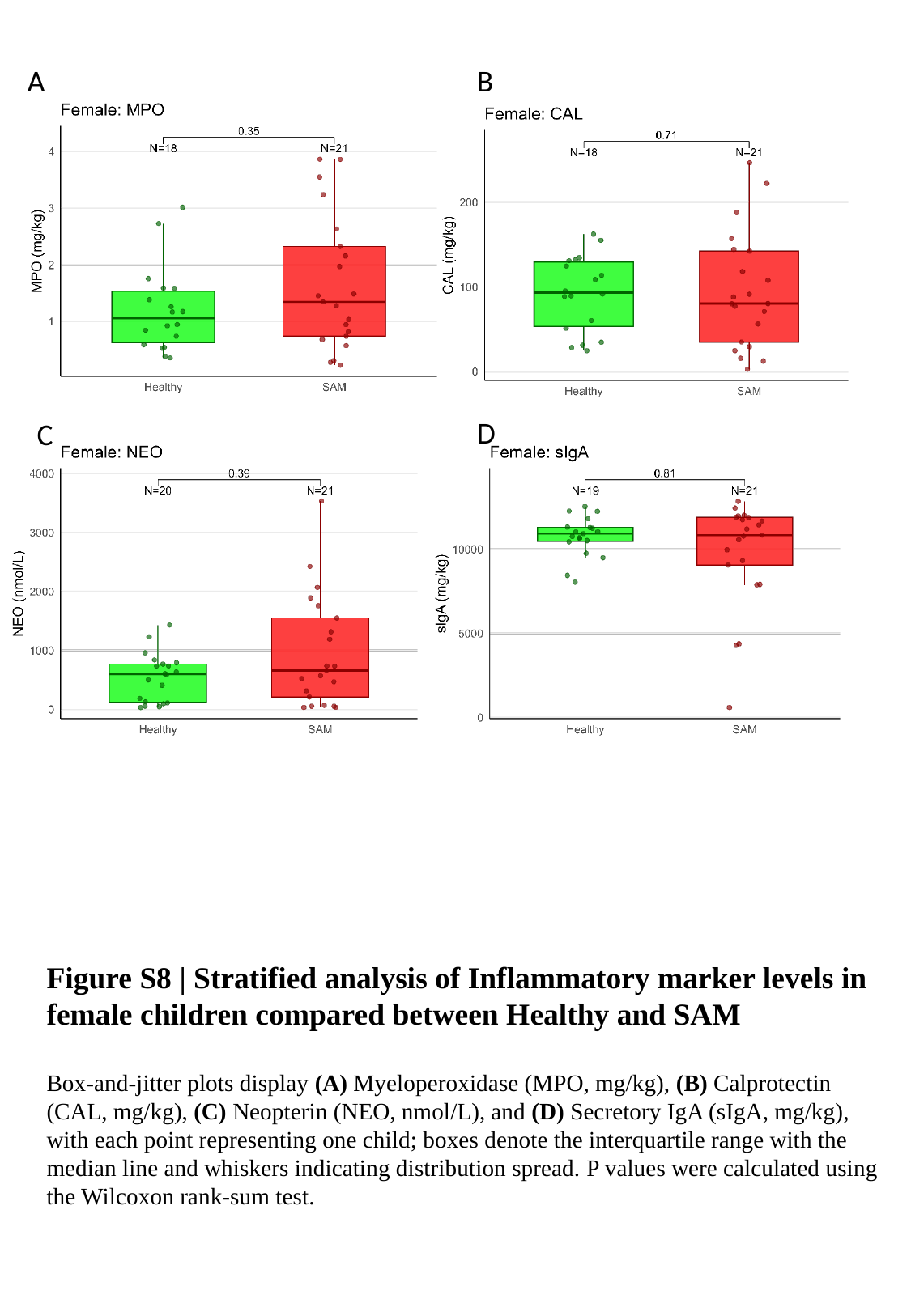

A
B
D
C
Figure S8 | Stratified analysis of Inflammatory marker levels in female children compared between Healthy and SAM
Box‑and‑jitter plots display (A) Myeloperoxidase (MPO, mg/kg), (B) Calprotectin (CAL, mg/kg), (C) Neopterin (NEO, nmol/L), and (D) Secretory IgA (sIgA, mg/kg), with each point representing one child; boxes denote the interquartile range with the median line and whiskers indicating distribution spread. P values were calculated using the Wilcoxon rank-sum test.

### Slide 9
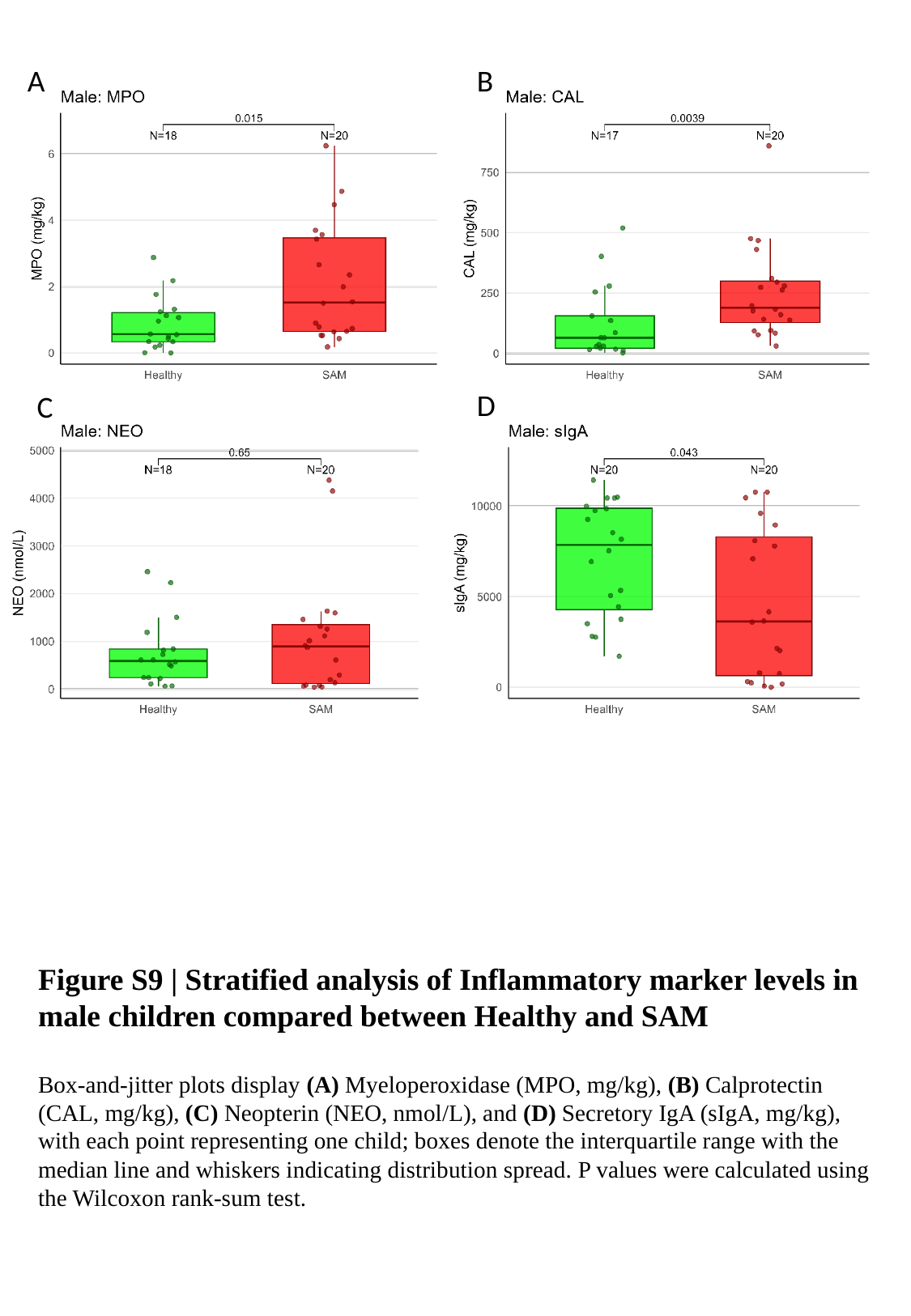

A
B
D
C
Figure S9 | Stratified analysis of Inflammatory marker levels in male children compared between Healthy and SAM
Box‑and‑jitter plots display (A) Myeloperoxidase (MPO, mg/kg), (B) Calprotectin (CAL, mg/kg), (C) Neopterin (NEO, nmol/L), and (D) Secretory IgA (sIgA, mg/kg), with each point representing one child; boxes denote the interquartile range with the median line and whiskers indicating distribution spread. P values were calculated using the Wilcoxon rank-sum test.

### Slide 10
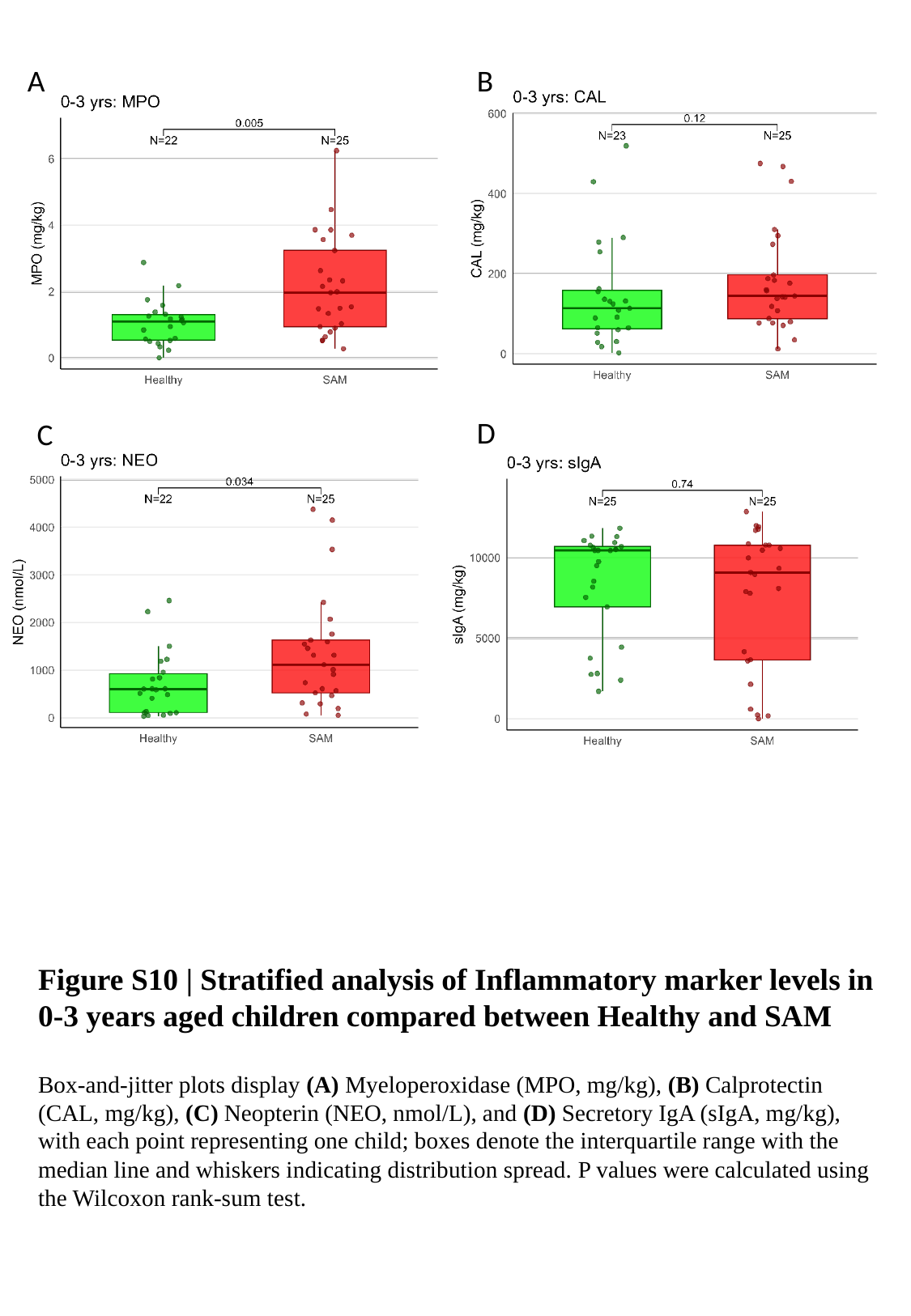

A
B
D
C
Figure S10 | Stratified analysis of Inflammatory marker levels in 0-3 years aged children compared between Healthy and SAM
Box‑and‑jitter plots display (A) Myeloperoxidase (MPO, mg/kg), (B) Calprotectin (CAL, mg/kg), (C) Neopterin (NEO, nmol/L), and (D) Secretory IgA (sIgA, mg/kg), with each point representing one child; boxes denote the interquartile range with the median line and whiskers indicating distribution spread. P values were calculated using the Wilcoxon rank-sum test.

### Slide 11
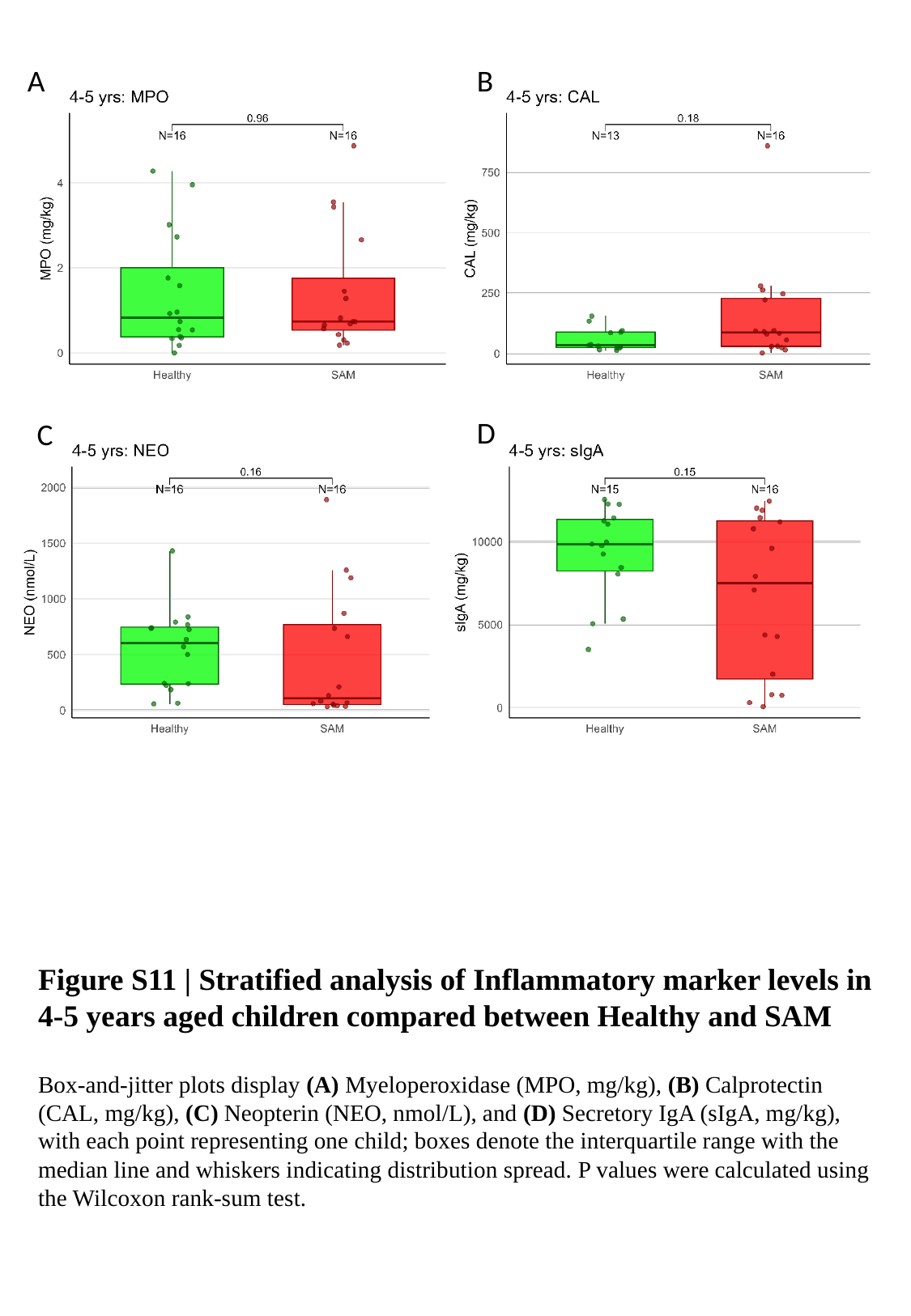

A
B
D
C
Figure S11 | Stratified analysis of Inflammatory marker levels in 4-5 years aged children compared between Healthy and SAM
Box‑and‑jitter plots display (A) Myeloperoxidase (MPO, mg/kg), (B) Calprotectin (CAL, mg/kg), (C) Neopterin (NEO, nmol/L), and (D) Secretory IgA (sIgA, mg/kg), with each point representing one child; boxes denote the interquartile range with the median line and whiskers indicating distribution spread. P values were calculated using the Wilcoxon rank-sum test.

### Slide 12
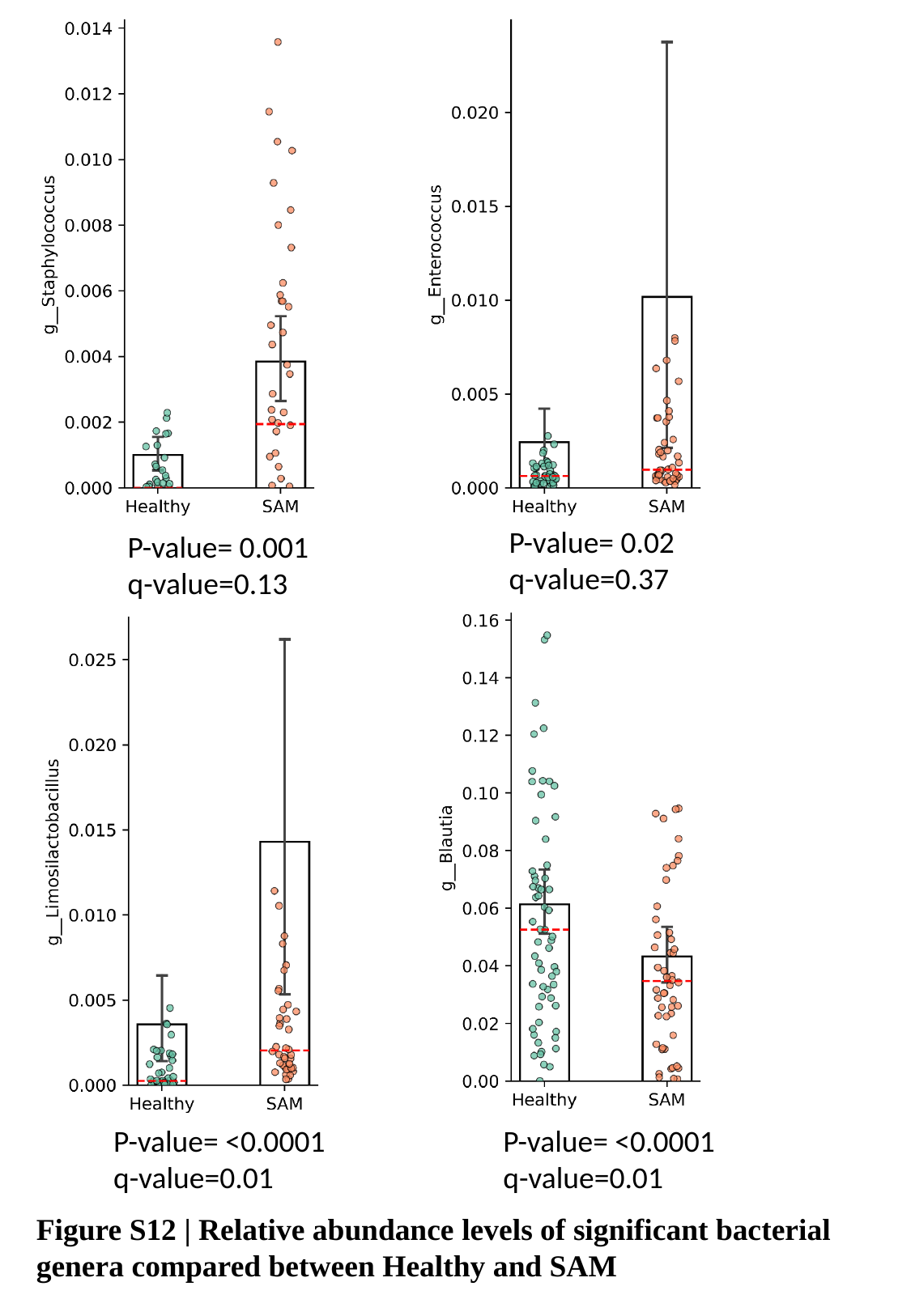

P-value= 0.02
q-value=0.37
P-value= 0.001
q-value=0.13
P-value= <0.0001
q-value=0.01
P-value= <0.0001
q-value=0.01
Figure S12 | Relative abundance levels of significant bacterial genera compared between Healthy and SAM

### Slide 13
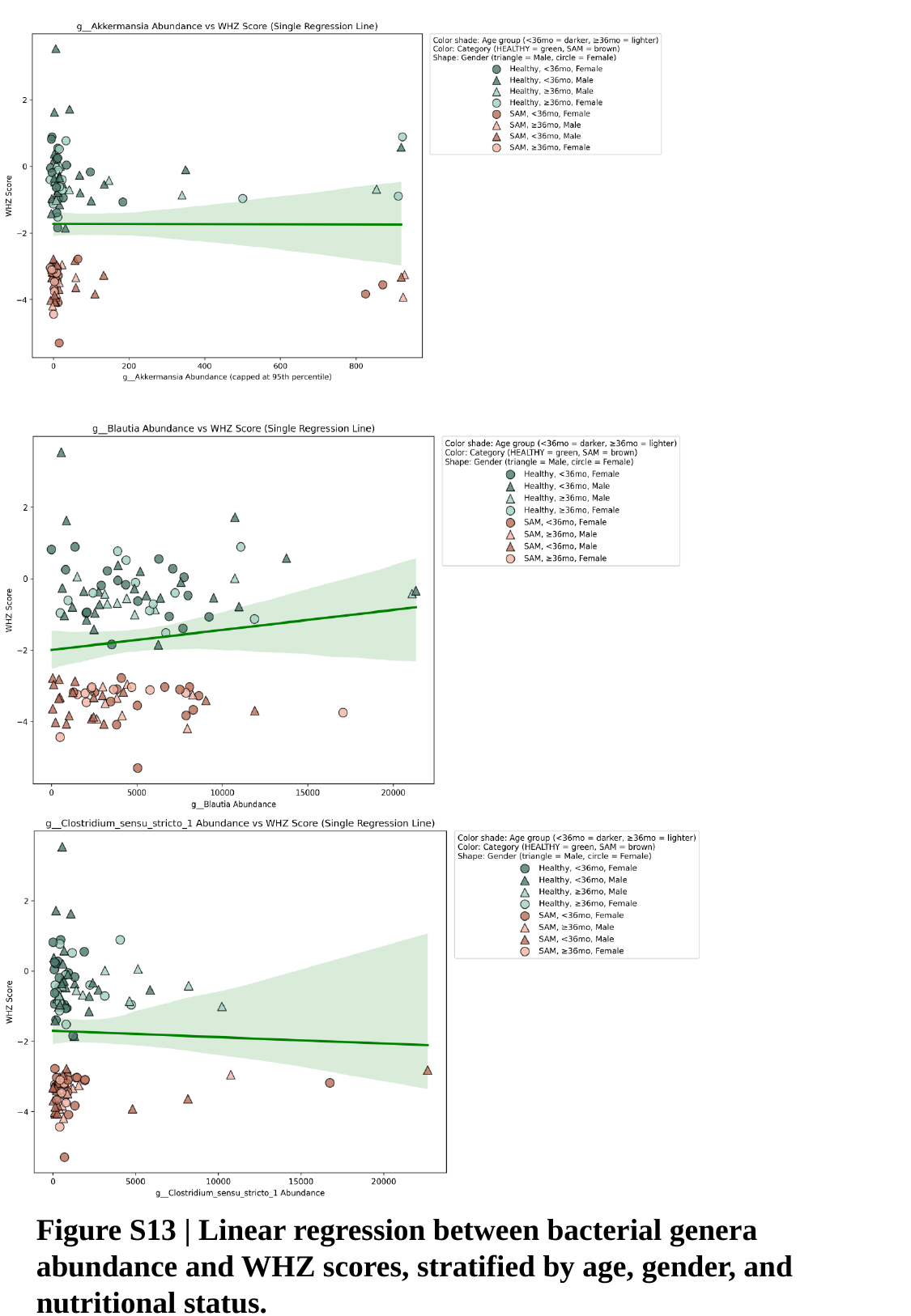

Figure S13 | Linear regression between bacterial genera abundance and WHZ scores, stratified by age, gender, and nutritional status.

### Slide 14
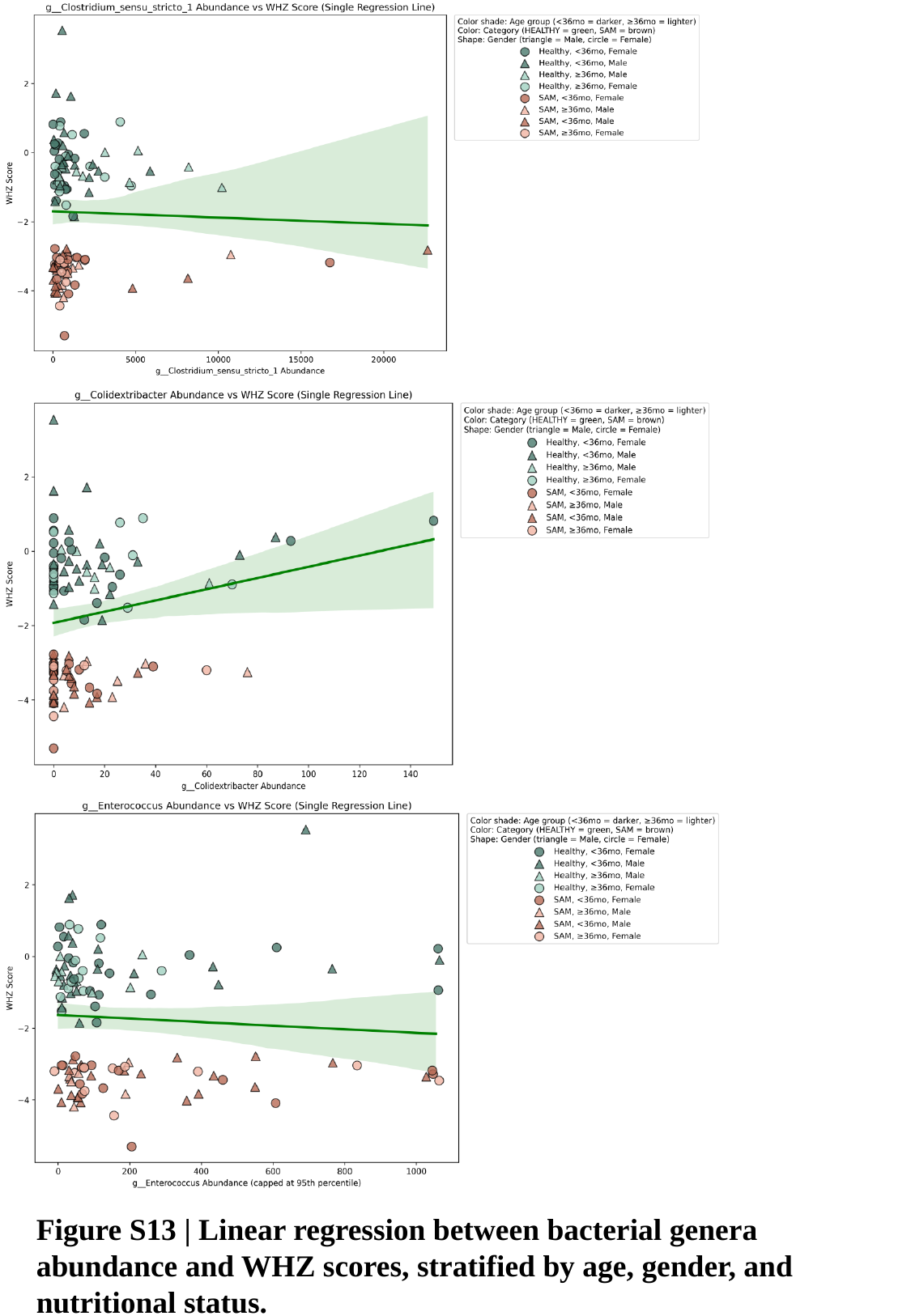

Figure S13 | Linear regression between bacterial genera abundance and WHZ scores, stratified by age, gender, and nutritional status.

### Slide 15
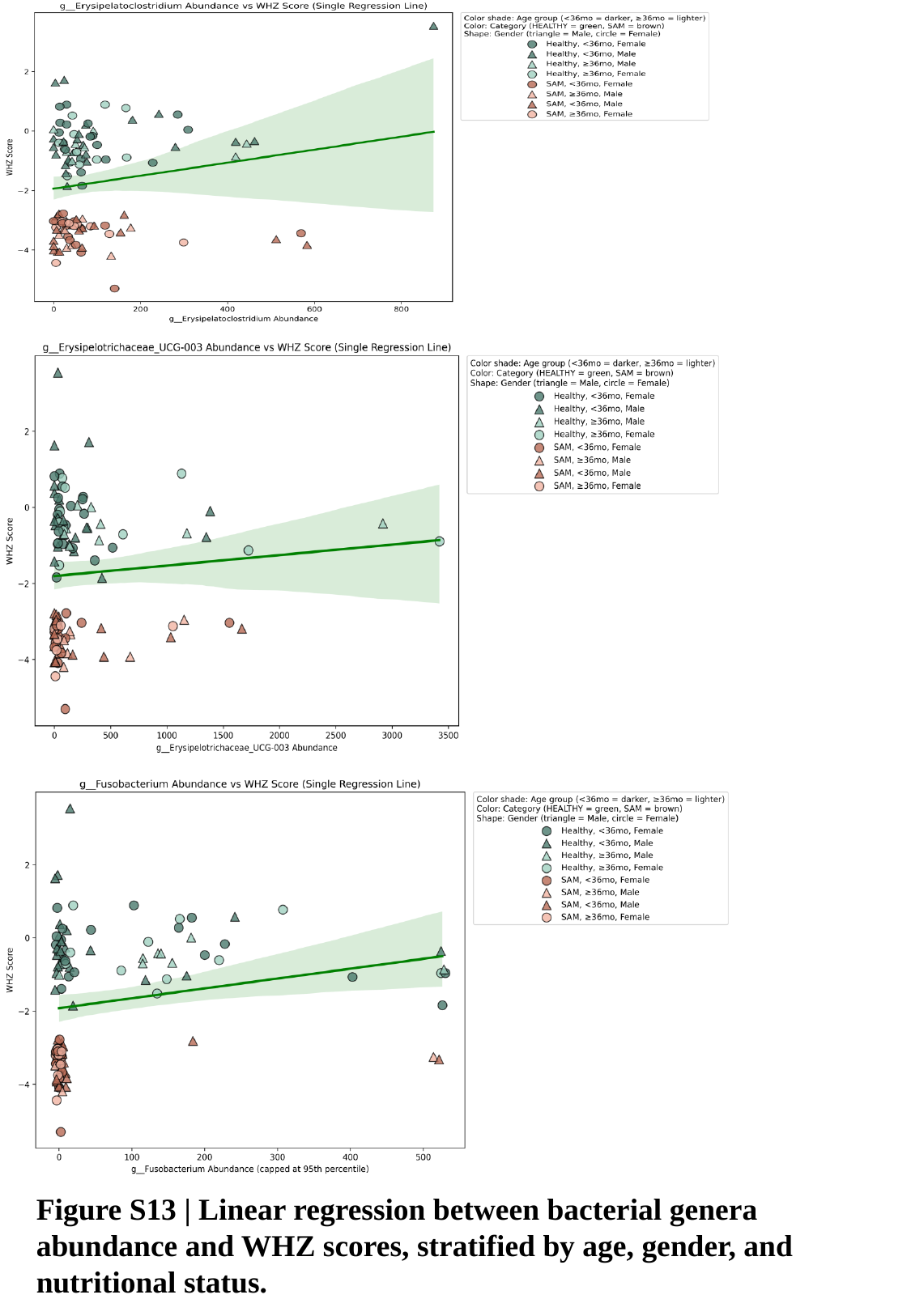

Figure S13 | Linear regression between bacterial genera abundance and WHZ scores, stratified by age, gender, and nutritional status.

### Slide 16
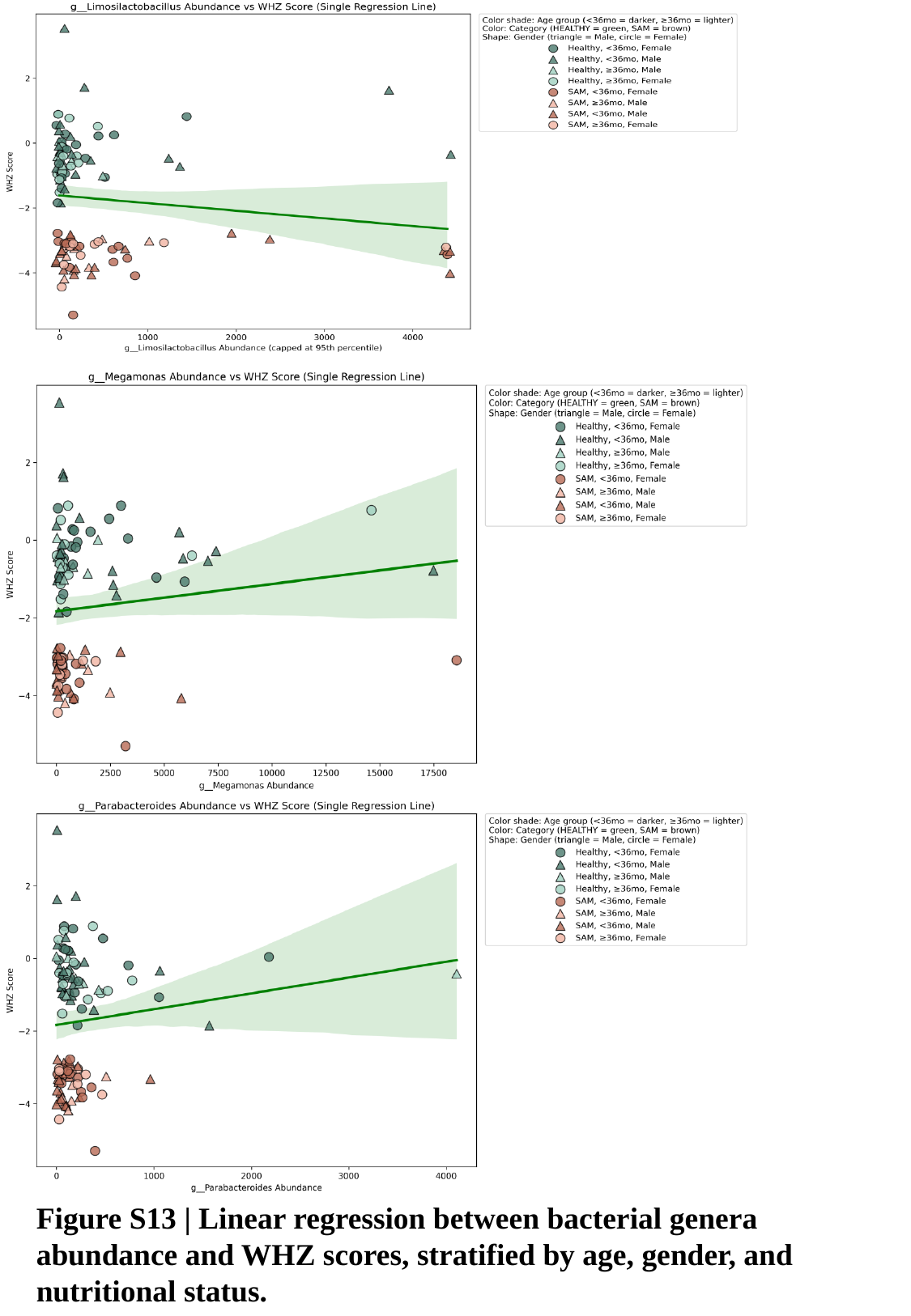

Figure S13 | Linear regression between bacterial genera abundance and WHZ scores, stratified by age, gender, and nutritional status.

### Slide 17
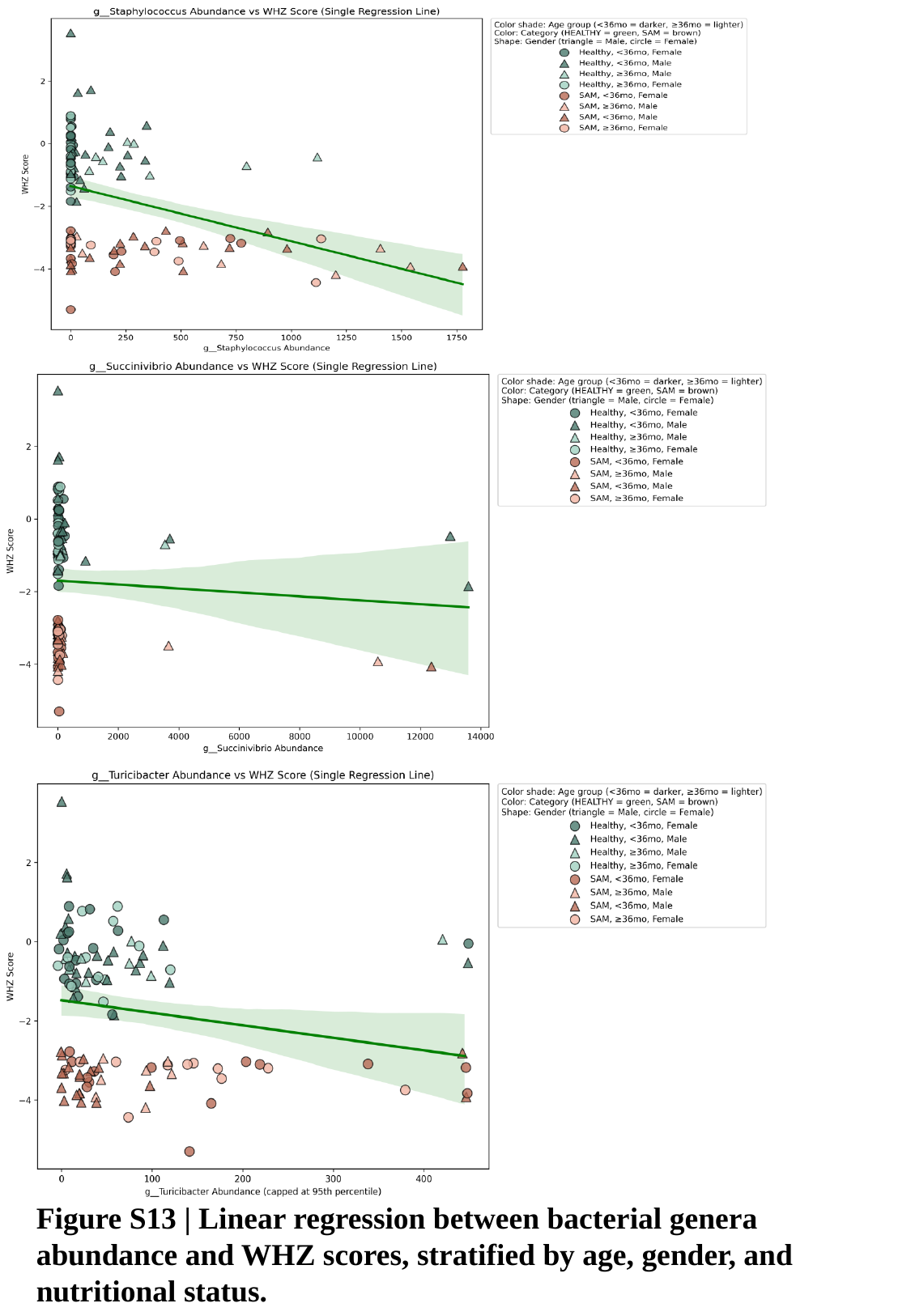

Figure S13 | Linear regression between bacterial genera abundance and WHZ scores, stratified by age, gender, and nutritional status.

### Slide 18
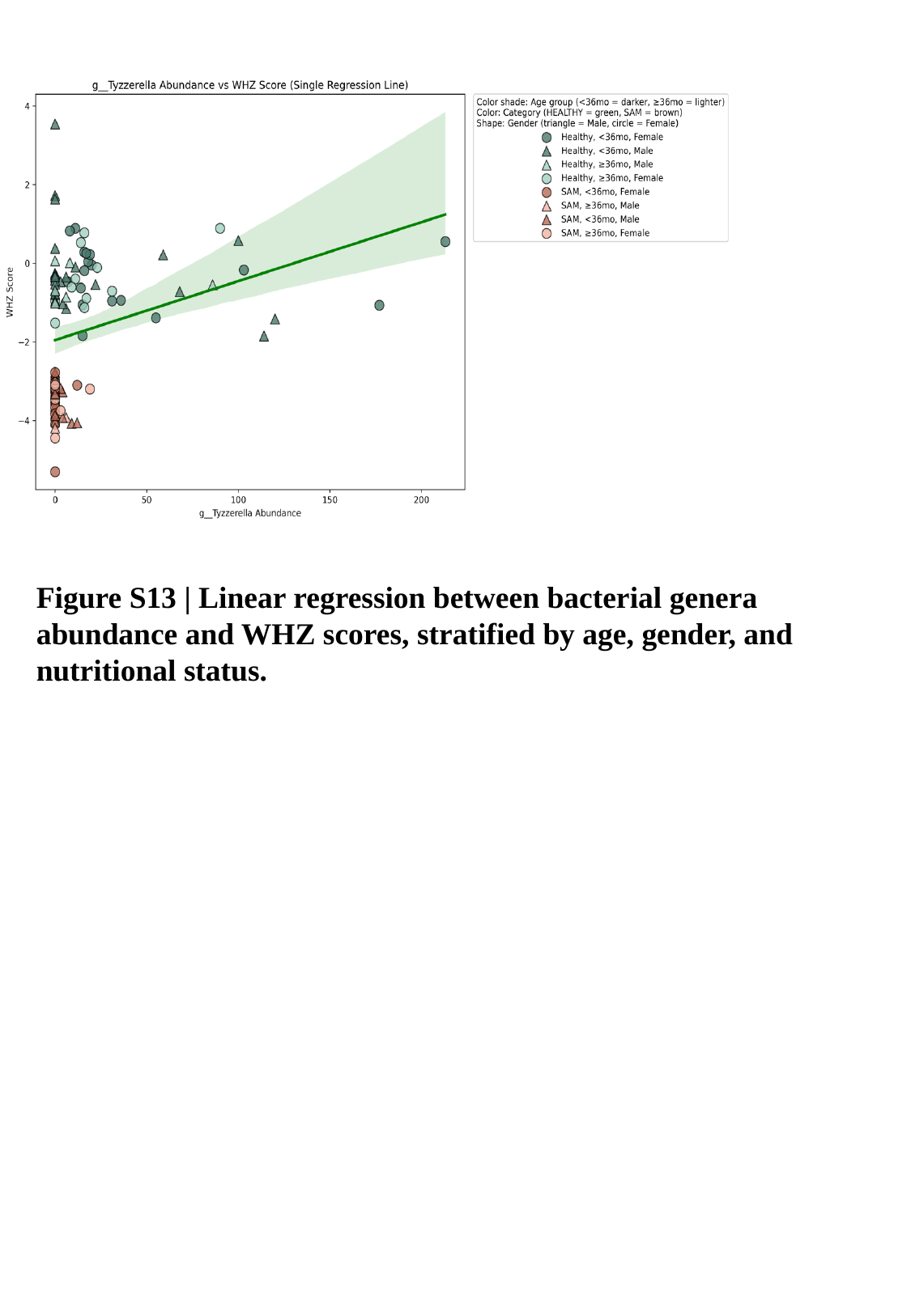

Figure S13 | Linear regression between bacterial genera abundance and WHZ scores, stratified by age, gender, and nutritional status.
