## Supplementary material for "Gut Microbial Dysbiosis and Elevated Mucosal Inflammation in Severe Acute Malnutrition: A Case-Control Study from India": Questionnaire for participant

Functional assessment of active and secretory IgA targeted and non-targeted bacterial groups in severe acute malnutrition (SAM)

Anganwadi Centre ID

Participant ID

*S - SAM; N - Healthy [S/NAWC-IDCHILD-ID]*

Date of SUBJECT enrolment

*Interview Date*

yyyy-mm-dd

Phone Number

Father's Name

Father's occupation

*Record exact job title*

Father's dietary choice

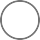
 Vegetarian

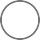
 Non-vegetarian

### Father's ailment

History of any major ailment of FATHER

*Select multiple incase of multiple ailments*

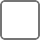
 Diabetes

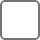
 Hypertension
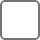
 Heart disease
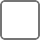
 Dyslipidaemia
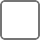
 Liver disease
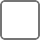
 Thyroid

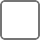
 None

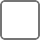

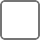
 Don't know/ not sure Other

Describe what 'Other' diseases he has

Father's height

*(in cms)*

Father's weight

*(in kgs)*

Mother's Name

Mother's occupation

*Record exact job title*

Mother's age

*(in Years)*

Mother's dietary choice

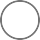
 Vegetarian

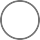
 Non-vegetarian

Mother's educational status

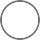
 No education

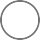
 Primary education

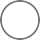
 Secondary education (10th)
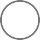
 Intermediate (12th)

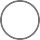
 Degree (College)

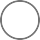

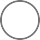
 Specialisation (Master's, Engineering etc) Don't know

### Mother's ailment

History of any major ailment of MOTHER

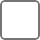
 Diabetes

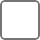
 Hypertension
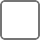
 Heart disease
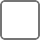
 Dyslipidaemia
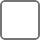
 Liver disease
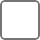
 Thyroid

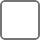
 None

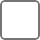
 Don't know/ not sure
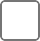
 Other

Describe what 'Other' diseases she has

Mother's Height

*(in cms)*

Mother's weight

*(in kgs)*

Consanguinity

*(Biologically related parents)*

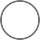
 Yes
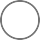
 No

Whether parents are vaccinated against COVID?

*(at least one dose)*

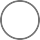
 Father
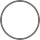
 Mother

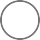
 Both parents
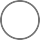
 None

Has any family member suffered from Covid or Covid like symptoms?

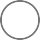

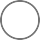
 Yes No

### Religion

What is the religion of the head of the household?

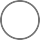
 HINDU
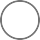
 MUSLIM
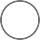
 CHRISTIAN
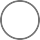
 SIKH

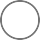
 BUDDHIST
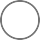
 JAIN

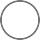
 JEWISH

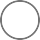
 PARSI/ZOROASTRIAN
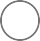
 NO RELIGION

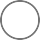
 OTHER

What religion does the head of the household follow (if response is 'OTHER')?

Caste/Tribe

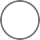
 General

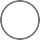
 Scheduled caste (SC)
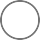
 Scheduled tribe (ST)

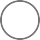
 Other backward class (OBC)
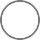
 None of them

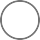
 Don't Know

Place of residence

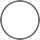
 City_Urban
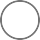
 City_Slum

Total number of people in the household

Where is the cooking done?

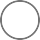
 Inside the House
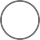
 Outdoors

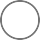

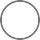
 In a separate building Other

Do you have a separate room which is used as a kitchen?

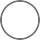
 Yes
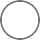
 No

How many rooms in this household are used for sleeping?

Source of drinking water
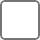
 Piped into dwelling
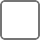
 Piped into plot/yard
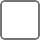
 Public tap/stand pipe
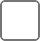
 Tubewell/Borehole
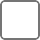
 Protected well

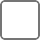
 Unprotected well
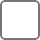
 Protected spring
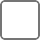
 Unprotected spring
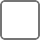
 Rainwater

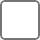
 Tanker truck

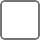
 Cart with small tank
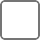
 Surface water

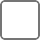
 Bottled water

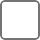
 Community water plant
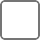
 others

Is Toilet facility available in the house

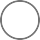
 Yes
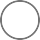
 No

Does this household have a BPL card?

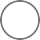

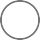
 Yes
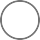
 No

Don't Know

Does your household have the following

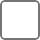
 Electricity
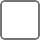
 Mattress

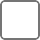
 Pressure cooker
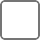
 Chair

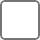
 Cot/Bed
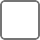
 Table

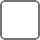
 Electric fan

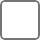
 Radio/Transistor
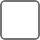
 TV

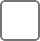
 Sewing machine
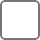
 Mobile

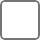
 Landline Telephone
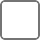
 Internet

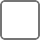
 Computer
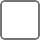
 Refrigerator
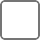
 AC/Cooler

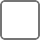
 Washing machine
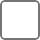
 Watch/Clock

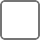
 Bicycle

Motorcyle/Scooter Animal-drawn cart Car

Water pump Thresher Tractor

Name of the Child

Date of birth of child

yyyy-mm-dd

Gender of the child

Male

Female

Does the child suffer from any chromosomal anomaly/cleft in lips?

Yes No

How many children does the mother have?

How many children are below 5 years of age?

Birth order of the enrolled child

Gestational time of the enrolled child

Full term Preterm Don't know

Place of delivery

Home Hospital Others

Mode of delivery

Vaginal Caesarian Forceps

Episiotomy (cut in the area between the vagina and anus during childbirth)

Birth weight of the child

Age of introduction of complimentary food

*(Food other than breast milk; age in months)*

Major complimentary food used

Rice based Wheat based Millet based Others

### Breast feeding

Is the child still breastfed?

Yes No

No. of breastfeeds per day

0-1

1-2

2-3

3-4

4-5

5-6

6-7

7-8

More than 8

Child's dietary choice

Vegetarian

Non-vegetarian

### Child Immunisation

Immunisation History of enrolled child

*Only fill data from 'Immunisation Card'*

Birth (BCG, OPV-0, HepB)

6 Weeks (OPV-1, Pentavalent-1, RVV-1, fIPV-1) 10 Weeks (OPV-2, Pentavalent-2, RVV-2)

14 Weeks (OPV-3, Pentavalent-3, fIPV-2, RVV-3) 9-12 Months (MR-1)

16-24 Months (MR-2, DPT-Booster-1, OPV-Booster) 5-6 Years (DPT-Booster-2)

Immunisation Card 'Not available'

Immunisation History if Immunisation Card 'Not available'

*Verbal response of mother to be confirmed by AWC teacher*

As per schedule

Missed immunisation dose

Washing of child's hand with soap after toilet

Yes No

### Recent Illness

History of illness in the last four weeks(if any)

*Check prescription*

Fever

Diarrhoea Cough/cold Malaria Dengue Other

None

Describe what illness the child had if response is 'Other' for 'History of illness'

### Medications taken

Whether child has consumed any antibiotics or probiotics in the last four weeks?

*Check prescription*

Yes No

List all medications taken by the enrolled child in the last 4 weeks

Has the enrolled child taken deworming medication in 2021?

Yes No

How many times in general does the child pass stool per day?

0-1

1-2

2-3

3-4

More than 4

Overall consistency of the child's stool

Firm to soft (easily passed) Hard (difficulty in passing) Semi-solid (runny)

Liquid (runny)

### Pica History

Does the child regularly craves or consumes non-food material (Pica History)

Yes No

List all non-food materials consumed by the enrolled child

### Hospitalisation History

Was the enrolled child ever hospitalised in the past?

Yes No

Details of why hospitalisation was required

*Check Health Card*

### Surgery History

Had the enrolled child have any surgery in past

Yes No

Details of all surgery performed on child and age when it was performed

Weight of child (in kgs)

Height of child (in cms)

MUAC of child (in cms)

CHILD: Triceps Skinfold Thickness(mm)(MM)

CHILD: Biceps Skinfold Thickness(mm)

CHILD: Suprailiac Skinfold Thickness(mm)

CHILD: Subscapular Skinfold Thickness(mm)

CHILD: Pulse

Child's body temperature

Does the Child have Anemia?

Yes No

Does the child have edema in both feet?

Yes No

Does the child show symptoms of jaundice?

Yes No

Does the child show signs of Hepatomegaly?

Yes No

Does the child show signs of Splenomegaly?

Yes No

CHILD: Abdominal examination
